# Explainable machine learning identifies predictors for postural instability and cognitive impairment in early Parkinson’s Disease

**DOI:** 10.64898/2026.09.30.26364120

**Authors:** Benjamin Sailer, Benjamin Roeben, Christoph Kiefer, Lars-Christian Achauer, Roswitha Kemmner, Isabel Wurster, Stefanie Lerche, Benjamin Bender, Tobias Lindig, Claudia Schulte, Christian Deuschle, Christian Erhardt, Thomas Gasser, Kathrin Brockmann, Oliver Kohlbacher, the DIFUTURE consortium, Stephanie Biergans, Inga Liepelt-Scarfone

## Abstract

Parkinson’s disease (PD) exhibits considerable variability in symptom onset, severity, and clinical trajectories, making prediction of individual disease progression a complex challenge. Leveraging large multimodal datasets from two independent cohorts of early PD patients, we examined predictive features for progression to mild cognitive impairment (MCI) and postural instability (PI) using Explainable Boosting Machines (EBM) - an interpretable machine learning (ML) approach. We found comparable predictive performances of EBM models using a minimal clinico-demographic dataset compared to an extended multimodal dataset including volumetric brain imaging and fluid biomarkers. Baseline age and Montréal Cognitive Assessment total score were identified as most important predictive features for both MCI and PI using the minimal dataset, while additional motor, cognitive and imaging features were identified using the extended dataset. Our study highlights the potential of ML predictions of cognitive and motor endpoints of PD progression at the individual level bringing personalized clinical decision-making within reach.

## Introduction

Onset and severity of clinical symptoms in Parkinson’s disease (PD) has found to be remarkably heterogeneous (1, 2). This observation has provided the basis for the data-driven identification and hypothesis-driven proposal of clinical PD subtypes (3–8), characterized by the occurrence or severity of varying motor and non-motor symptoms. Depending on the predominant symptoms, disease progression of PD patients differs (3, 4, 9–11). As a consequence, the time to reach critical disease milestones, such as motor (e.g. postural instability increasing patients fall risk) and non-motor complications (e.g. cognitive impairment), is variable among patients (1, 12, 13). Importantly, a combination of motor and non-motor complications is associated with a more rapid disease progression (11, 14–16), highlighting the importance to monitor the onset and severity of motor and non-motor symptoms already in early stages of the disease.

However, previous subtyping approaches currently employed for predicting PD progression lack clinical applicability and have significant methodologic shortcomings (6, 17). Thus, the ultimate goal of predicting individual disease progression in PD remains a complex challenge that is still not achieved yet. Integrating larger datasets of multimodal data and application of novel statistical algorithms promises to enable identification of biomarkers for more accurate prediction of disease progression and related milestones, but in turn comes with even higher complexity of data analysis. Approaching this challenge, recent advances in machine learning (ML) methods promise to deliver more robust and accurate predictions of progression even in early disease stages. Several studies have employed various ML algorithms to investigate different aspects of PD. By leveraging large multimodal datasets including lifestyle, demographic, clinical, imaging and digital motor data as well as genetic and fluid biomarkers machine learning has been applied for predicting clinical as well as prodromal PD diagnosis (18–20), clinical subtypes, associated disease progression and clinical endpoints (21–23).

To date, only few studies have focused on specific motor milestones of PD, e. g. ML-based prediction of freezing of gait based on structural brain volumetric data compared to clinical and fluid biomarker data (24). Furthermore, knowledge regarding data-driven predictors of postural instability (PI) in PD is still scarce. Early onset of PI in PD indicates a more rapid disease progression (25–27), potentially within the frame of a differential diagnosis (28, 29). More importantly, PI has negative impact on gait and walking performance associated with increased risk and fear of falling significantly affecting quality of live (30). Therefore, identification of persons at risk for PI is essential to optimize patient’s treatment (31). Such early detection opens the window for early pharmaceutical and physical interventions.

Cognitive impairment increases the risk for dementia, nursing home admission and mortality, dramatically affecting patient’s and caregiver quality of life (32). Due to the fact that cognitive impairment is an important milestone in PD, a larger number of studies has focused on ML-based prediction of cognitive decline in PD using multimodal biomarker, clinical and imaging data (33–38). Most of these studies used large datasets to discover predictors for cognitive impairment in early stage PD (18, 34–36, 39–44) resulting in different combinations of demographic, motor and non-motor predictors as well as imaging and fluid biomarkers.

However, only few studies applied ML to data similar to that collected in routine clinical practice (45). This raises the question, whether a dataset reflecting data routinely collected in a clinical care setting, where time for patient assessment is limited, has the same predictive value compared to larger research-based multimodal datasets, e.g. including imaging data, which is more expensive and not necessarily available in primary care.

Therefore, in the present study we set out to predict PD disease progression as assessed by two predefined clinical disease milestones - mild cognitive impairment (MCI) and PI. We investigated features contributing to these two clinical disease milestones/endpoints by employing a ML algorithm –EBM – that facilitates explainability of the ML model. To this end, we leveraged multimodal datasets including clinical, demographic, genetic, fluid biomarker and imaging data from two large independent cohorts of patients in early PD stages: the Tuebingen Parkinson Cohort (TuePaC) and the Parkinson’s Progressive Markers Initiative (PPMI) study cohort. We compared predictive performances of EBM models in three subsets of the datasets: (i) a minimal data set encompassing only clinical and demographic variables frequently assessed in routine clinical care of PD patients; (ii) an extended data set additionally including genetic and fluid biomarkers; and (iii) a sub-dataset including volumetric brain imaging data in a nested cohort of the PPMI study.

## Results

For analysis of the cognitive endpoint MCI, data from 334 patients of the PPMI cohort and 213 patients of the TuePaC cohort, and for analysis of the motor endpoint PI, data from 339 patients of the PPMI cohort and 334 patients of the TuePaC cohort were available (see Figure 1). For the PPMI imaging subcohort, data from 209 patients were available for the MCI endpoint and from 210 patients for the PI endpoint (see Figure 1).

**Figure 1:**
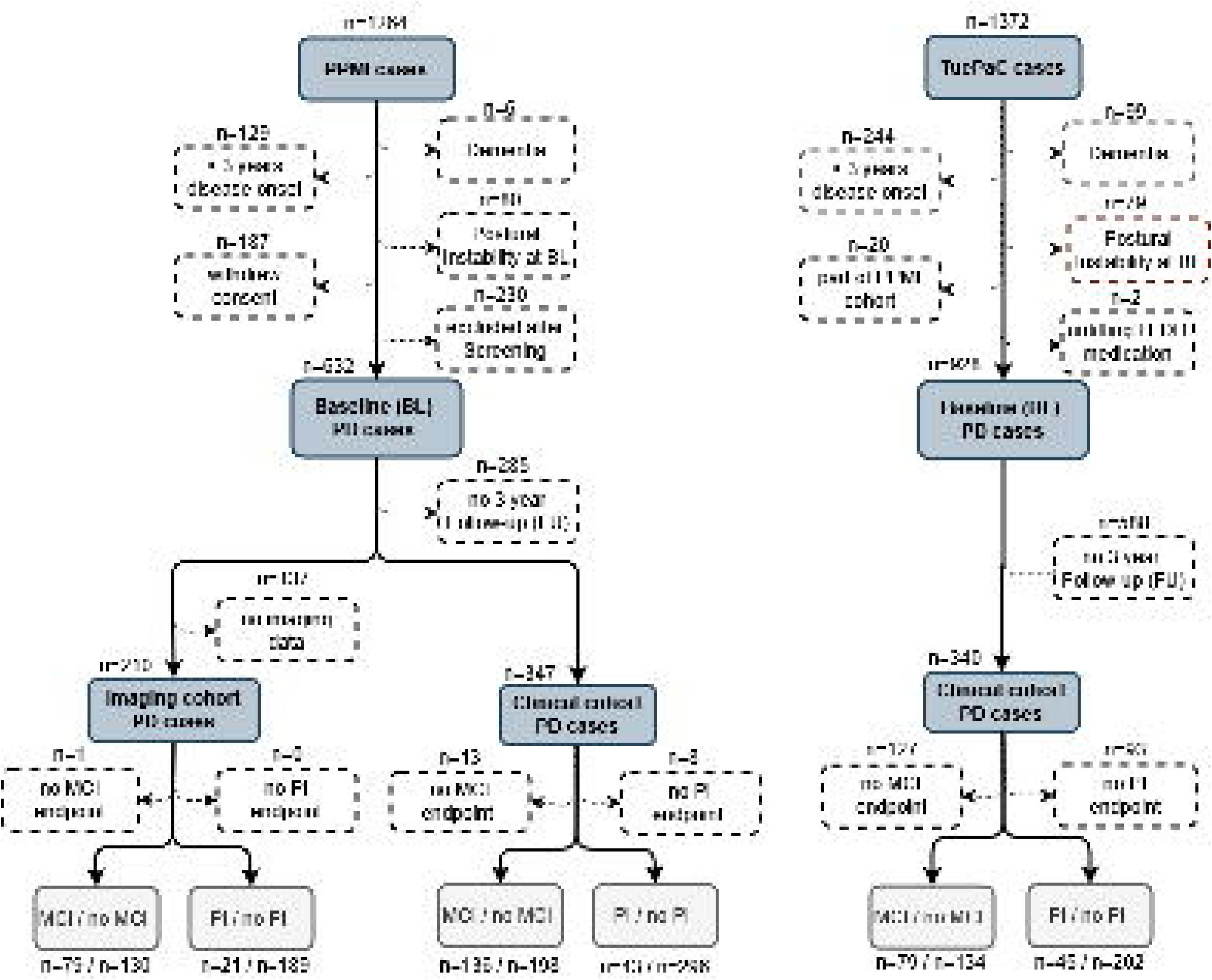
Flowchart of the case selection process of the clinical and imaging datasets of the Tuebingen Parkinson Cohort (TuePaC) and the PPMI study cohort. Resulting absolute numbers of the predefined clinical endpoints Mild Cognitive impairment (MCI) and postural instability (PI) at 3-year follow-up in the respective cohort datasets are presented at the bottom of the flowchart.

### Clinical and demographic characteristics of the TuePaC and PPMI cohorts

Details on the clinical and demographic characteristics, and frequencies of the clinical endpoints for MCI and PI in the TuePaC and PPMI cohorts are shown in Table 1.

**Table 1:**
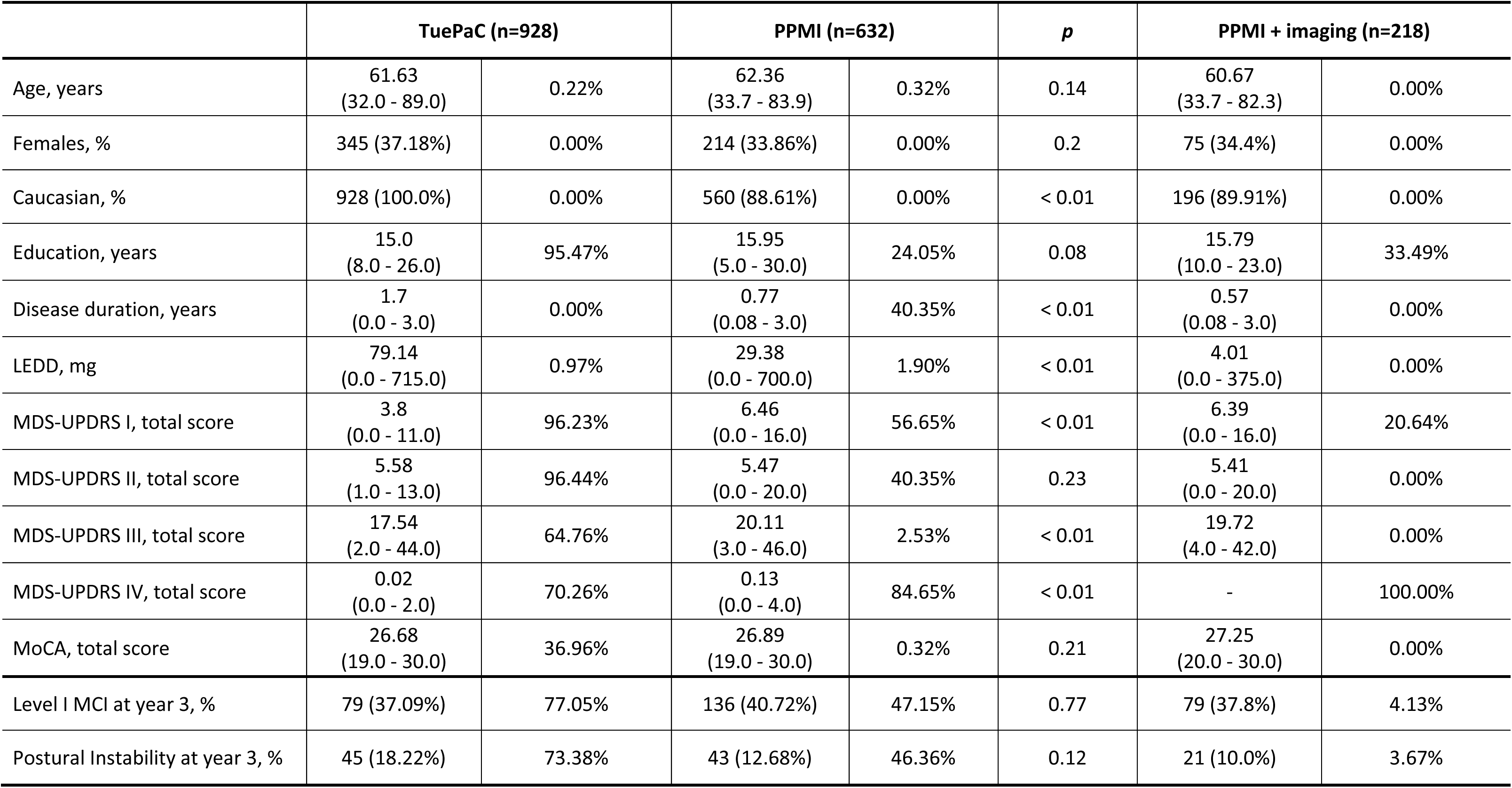
Clinical and demographic characteristics, and frequencies of the clinical endpoints MCI and PI in the TuePaC and PPMI cohorts. Data is presented as mean with min and max in brackets or frequency with percentage in brackets. Mann-Whitney-U test was used for continuous data and χ^2^ test was used for categorial data. Two-sided p < 0.05 is presented in bold. Percentage of missing data per variable is shown in a separate column.

The TuePaC cohort had more individuals of Caucasian descent compared to the PPMI cohort (TuePaC: 100% Caucasians vs. PPMI: 88.61% Caucasians; *p*<0.01). Disease duration (TuePaC 1.70±0.73 years vs. PPMI 0.77±0.73 years; *p*<0.01) and LEDD (TuePaC 79.14±137.95 mg vs. PPMI 29.38±93.94 mg; *p*<0.01) were higher in the TuePaC cohort, whereas in contrast scores of the MDS-UPDRS I (TuePaC 3.80±3.18 points vs. PPMI 6.46±3.90 points; *p*<0.01), MDS-UPDRS III (TuePaC 17.54±7.62 points vs. PPMI 20.11±8.88 points; *p*<0.01) and MDS-UPDRS IV (TuePaC 0.02±0.16 points vs. PPMI 0.13±0.55 points; *p*<0.01) were higher in the PPMI cohort. No differences were found for gender or for MDS-UPDRS II and MoCA scores between groups (*p*>0.05).

The frequencies of the two predefined clinical endpoints MCI and PI were comparable in both cohorts and all 3 PPMI subsets. At the 3-year follow-up, the clinical cognitive endpoint MCI was observed in 37.09% of the TuePac cohort and 40.72% of the PPMI cohort (*p*=0.99), while the clinical motor endpoint PI was reached by 18.22% in the TuePaC and 12.68% in the PPMI cohort (*p*=0.75).

### Prediction of clinical endpoints using Explainable Boosting Machine (EBM)

#### Model accuracies for prediction of MCI

For the prediction of the cognitive endpoint MCI, the EBM model based on the clinicodemographic minimal dataset yielded a moderate balanced accuracy of 0.62 and a receiver operating characteristics area under the curve (ROC-AUC) of 0.68 for MCI resulting in a sensitivity of 0.61 and a specificity of 0.63 (Table 2). Using the multimodal extended dataset, the performance of the EBM model showed lower ROC-AUC for MCI (minimal 0.68 vs extended 0.62; Table 2) as compared to the EBM model using the clinicodemographic minimal dataset, while balanced accuracies were identical. Furthermore, sensitivity of the model was slightly higher (minimal 0.61 vs extended 0.64; Table 2), but specificity was lower (minimal 0.63 vs extended 0.60; Table 2; Supplementary Table 3a & 4a).

**Table 2:** EBM performances of the clinicodemographic minimal, the multimodal extended and the imaging datasets for the prediction of the clinical endpoints MCI and PI. TP: True Positive, TN: True Negative, FP: False Positive, FN: False Negative.

| Performance Measures | Mild Cognitive Impairment (MCI) |  |  |  | Postural instability (PI) |  |  |  |
| --- | --- | --- | --- | --- | --- | --- | --- | --- |
|  | minimal | extended | PPMI + imaging | PPMI - imaging | minimal | extended | PPMI + imaging | PPMI - imaging |
| **Bal. Accuracy** | 0.62 | 0.62 | 0.68 | 0.65 | 0.62 | 0.62 | 0.63 | 0.59 |
| **ROC AUC-value** | 0.68 | 0.62 | 0.74 | 0.73 | 0.62 | 0.66 | 0.67 | 0.64 |
| **F1-Score** | 0.54 | 0.5 | 0.57 | 0.53 | 0.37 | 0.33 | 0.34 | 0.25 |
| **TP/TN/FP/FN** | 48/84/50/31 | 16/34/23/9 | 10/16/12/2 | 8/24/8/6 | 29/120/82/16 | 15/43/57/4 | 2/25/5/3 | 2/26/7/3 |
| **Precision** | 0.49 | 0.41 | 0.45 | 0.54 | 0.26 | 0.2 | 0.29 | 0.2 |
| **Sensitivity** | 0.61 | 0.64 | 0.79 | 0.56 | 0.64 | 0.79 | 0.44 | 0.38 |
| **Specificity** | 0.63 | 0.6 | 0.58 | 0.75 | 0.59 | 0.43 | 0.82 | 0.79 |
| **Youden Index** | 0.24 | 0.24 | 0.37 | 0.31 | 0.24 | 0.22 | 0.26 | 0.17 |

Integrating imaging features from brain volumetric data together with the clinico-demographic minimal dataset into the EBM model yielded slightly higher ROC-AUC for prediction of MCI (0.74 with imaging features vs 0.73 without imaging features; Table 2) and higher balanced accuracy compared to the EBM models using the minimal dataset without imaging features (0.68 with imaging features vs 0.63 without imaging features; Table 2; Supplementary Table 5a).

#### Model accuracies for prediction of PI

For the motor endpoint PI, the EBM model using the clinico-demographic minimal dataset showed a ROC-AUC of 0.62 with sensitivity of 0.64 and specificity of 0.59, and likewise moderate balanced accuracy of 0.62 (Table 2). In contrast to the results for MCI, the EBM model using the multimodal extended dataset showed higher ROC-AUC for prediction of PI (minimal 0.62 vs extended 0.66) as compared to the EBM model using the clinicodemographic minimal dataset. Again, sensitivity was higher (minimal 0.64 vs extended 0.79), at the expense of lower specificity (minimal 0.59 vs extended 0.43; Supplementary Table 3b & 4b).

Similarly to the EBM models for prediction of MCI, the EBM model combining the clinico-demographic dataset and imaging features resulted in higher balanced accuracy and higher ROC-AUC for prediction of PI compared to using the minimal dataset without imaging features (ROC-AUC: 0.67 with imaging features vs 0.64 without imaging features; balanced accuracy: 0.63 with imaging features vs 0.59 without imaging features; Table 2; Supplementary Tables 5b).

#### Predictive features for conversion to MCI

The most important features for prediction of conversion to MCI using the minimal dataset were the MoCA score and age followed by gender and PD family history (Figure 2). With regard to the direction of the effect of the two most important features, shape functions showed that lower MoCA scores and higher age predicted conversion to MCI (Figure 2 C & D). Furthermore, male gender was also predictive for MCI.

**Figure 2:**
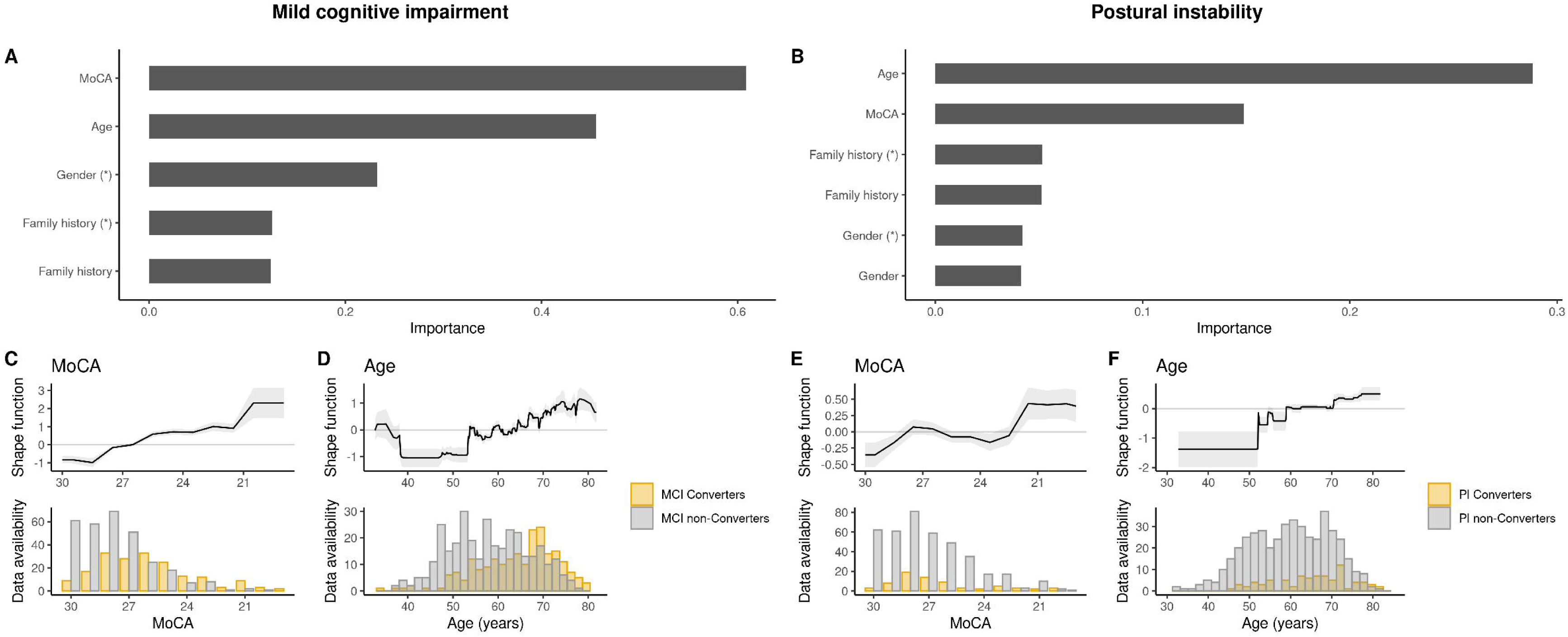
EBM prediction of clinical endpoints Mild Cognitive Impairment (MCI) and postural instability (PI) based on a clinicodemographic minimal dataset. EBM prediction model showing importance of risk factors predicting the predefined clinical endpoints MCI (A) and PI (B). Shape functions and data distribution for the two most important features MoCA and age are shown for MCI (C & D) and PI (E & F) respectively. Asterisks (*) indicate target encoded features.

The EBM model using the extended multimodal dataset verified the MoCA total score, as well as the MoCA memory and cube subscores, age and retirement status as the top 5 most important features for later onset of MCI (Figure 3A).

**Figure 3:**
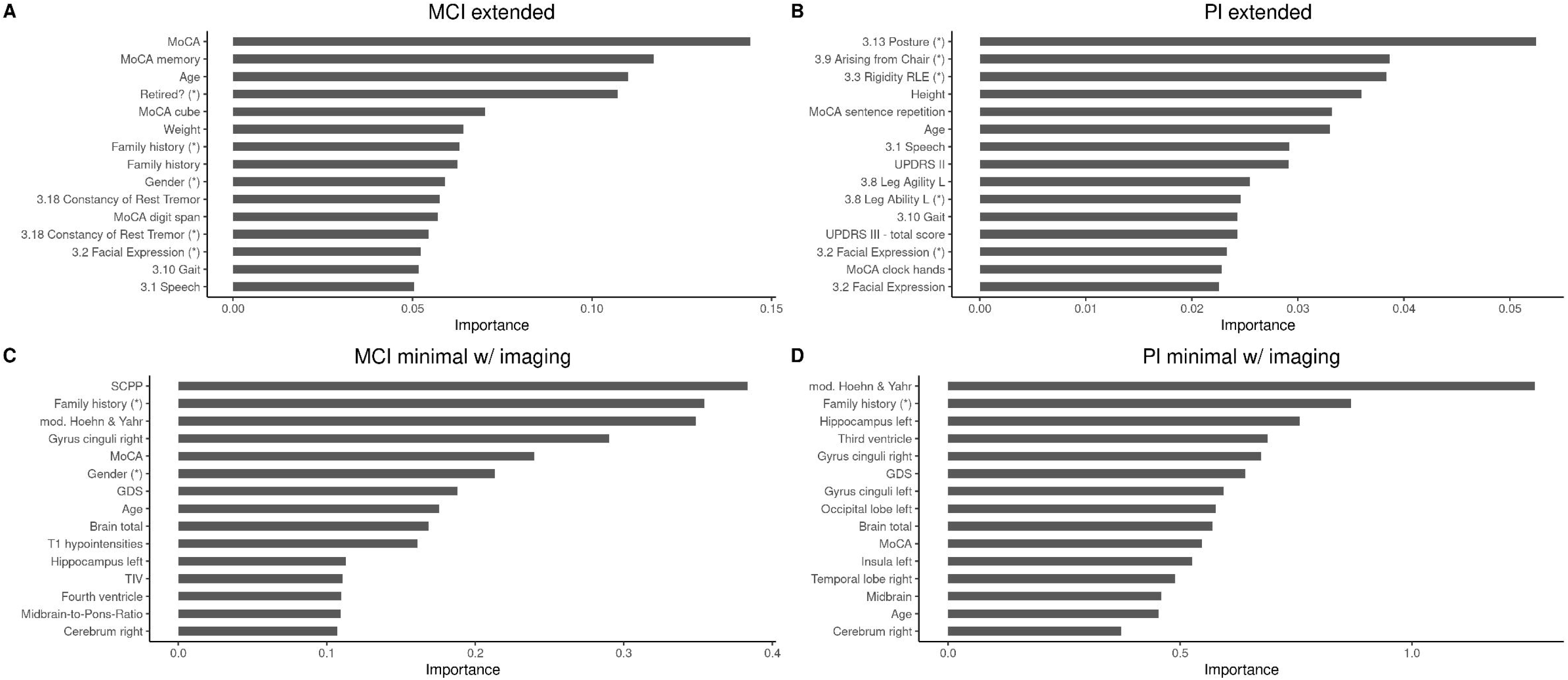
Feature importance from EBM models using an extended dataset for prediction of the clinical endpoints MCI (A) and PI (B). Asterisks (*) indicate target encoded features.

Combining the minimal clinico-demographic dataset and imaging features for prediction of MCI, the EBM model top-ranked several volumetric features with volume of the superior cerebellar peduncles (SCP) as the top feature, and volumes of the right gyrus cinguli and the left hippocampus and T1 hypointensities (Figure 4 A-D) as well as brain total volume in addition to clinical and demographic features in the feature importance ranking (e.g. family history of PD, modified H&Y, MoCA, gender, GDS and age; see Figure 3C).

**Figure 4:**
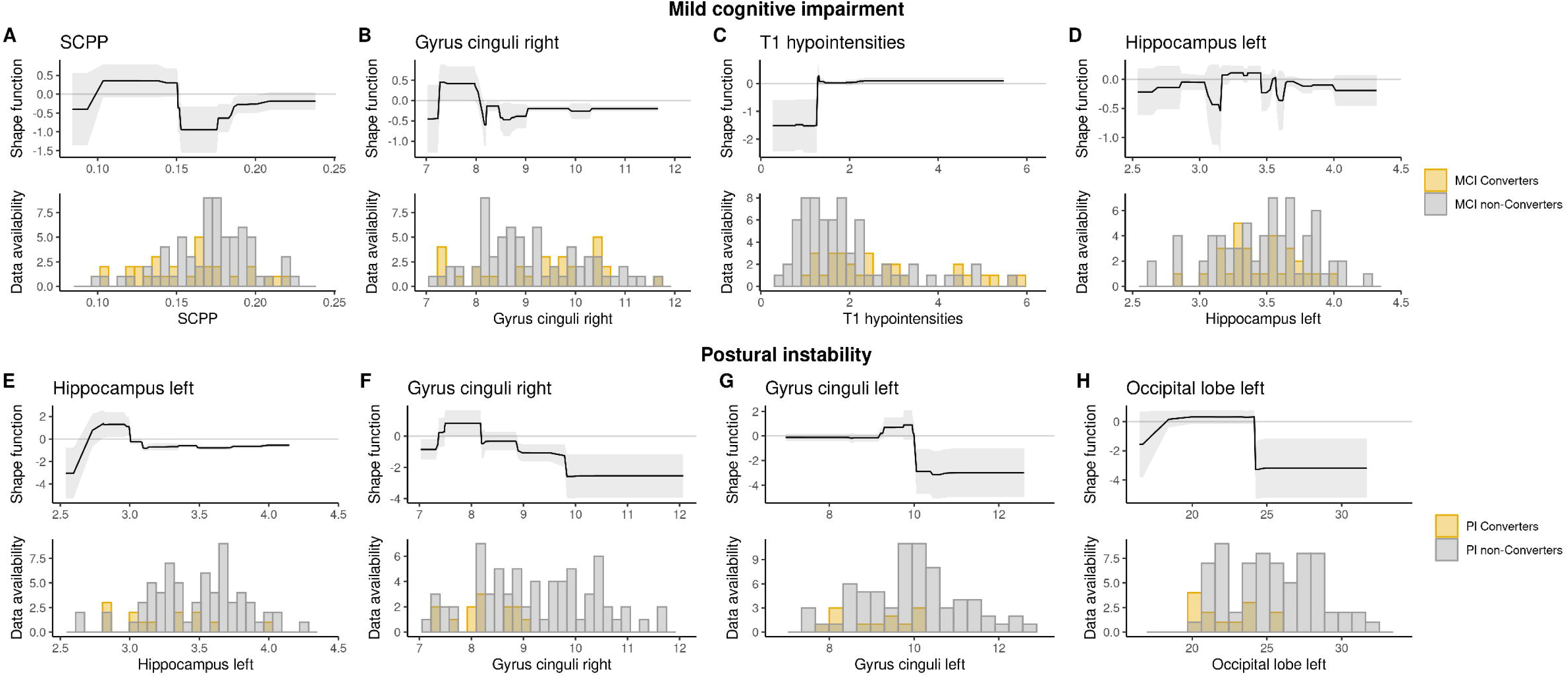
Volumetric features resulting from EBM models integrating imaging data for prediction of the cognitive and motor endpoints MCI and PI (E-H). Shape functions and data distribution for the most important imaging features are shown for MCI (A-D) and PI (E-H).

#### Predictive features for PI

For prediction of PI, the most important features using the minimal dataset were largely similar to those for the prediction of MCI (age, MoCA, PD family history and gender), however the order of importance of each single feature differed between the models (see Figure 2). Furthermore, lower MoCA scores and higher age predicted conversion to PI (Figure 2 E & F). Compared to this, the MDS-UPDRS III subitems “posture”, “arising from chair” and “rigidity of the right lower extremity” as well as height and the MoCA sentence repetition subscore were the top 5 most important features in the EBM model using the extended multimodal dataset (Figure 3B).

Finally, the most important imaging features for predicting conversion to PI included the volume of the left hippocampus, right and left gyrus cinguli, left occipital lobe (Figure 4 E-H) as well as volumes of the third ventricle and brain total volume (Figure 4) besides similar clinical and demographic features as compared to the cognitive endpoint MCI (Figure 3D).

## Discussion

In this study, we used an explainable ML algorithm – EBM – to determine which features contribute to the progression of two important clinical outcomes: cognitive decline indicated by the level I diagnosis of MCI, and PI as marker for motor worsening. In accordance with evidence from previous studies on the prediction of MCI, we provide supportive evidence improving the interpretability and generalizability of previous studies as well as novel insights into factors contributing to progression to the motor endpoint PI. To our knowledge, this is the first ML-based evaluation of PD progression with PI as a clinical motor endpoint.

We show that the performance metrics of generated EBM models for predicting progression to the two predefined clinical endpoints were largely comparable when using a minimal clinico-demographic dataset as compared to an extended multimodal dataset. Futhermore, by integrating features from automated brain volumetry resulting in slight additional improvement of predictive performance, we show that easily accessible imaging features provide an added value promising to enable a more precise, individualized characterization of factors contributing to PD progression. Thus, our study supports the value of data collected in routine clinical care to identify those patients at high risk for rapid progression. We approached this by defining a core clinico-demographic minimal dataset encompassing data from assessments that are already widely implemented in the real-world clinical care of PD patients.

Unlike other ML algorithms, EBMs are a class of generalized additive models (GAMs) that are inherently interpretable. In order to take advantage of this benefit and improve the clinical applicability of such ML algorithms in real-world settings, the selection of input features and the number of features selected are of critical relevance.

Using the minimal clinico-demographic dataset, we show that the predictive performance metrics of the trained EBM models are generally comparable to those obtained with an extended multimodal dataset. Furthermore, EBM models using both datasets delivered similar results in the local TuePaC cohort compared to the for the feature selection.

As automated pipelines for processing of imaging data become increasingly available (46, 47), we also evaluated the added value of integrating volumetric imaging features for prediction of the two clinical endpoints. We found an overall slight increase of predictive performance metrics when combining the minimal clinico-demographic dataset and volumetric imaging features for prediction of conversion to both MCI and PI.

These findings exemplify that datasets of core clinico-demographic data readily enable ML-based prediction of disease progression to MCI and PI even in the early phase of PD. Notably, we found the same predictors in the local, clincial TuePaC cohort as compared to the multisite comprehensive PPMI study. More extensive multimodal datasets including easily accessible imaging data might facilitate more detailed and in-depth phenotypig of PD progression and its contributing factors, which might guide future research into PD progression and aid in conceptualizing more personalized treatments for PD patients.

The majority of previous studies involving PD patients using ML methods compared performances of different algorithms and used *“SHapley Additive exPlanations”* (SHAP) (48) to generate model insights (18, 34, 35, 49, 50). However, SHAP have several limitations that can lead to misleading interpretations of feature importance, such as assigning non-zero importance to irrelevant features or ranking irrelevant features above relevant ones (51, 52). These limitations may affect the trustworthiness and clinical usefulness of SHAP explanations in biomedical contexts.

Alternative Explainable AI approaches such as Explainable Boosting Machines (EBM) might provide more robust and clinically actionable insights, fostering a deeper understanding of the underlying biomedical processes (53–57). EBMs enable interpretability of data by generating predictions based on the summation of average risk scores associated with each features’ value, along with baseline bias terms for each individual data point (58). The resulting *‘shape functions’* for each feature provide a transparent understanding of the models’ behavior and the underlying data structure. However, it is important to note that these shape functions represent associations (correlations) rather than causal relationships.

With regard to the cognitive endpoint conversion to MCI, feature importance ranking and evaluation of the respective shape functions of the EBM model using the minimal dataset showed that lower MoCA scores and higher age predicted MCI in line with numerous previous ML-based studies (35, 41, 43, 44). Furthermore, the EBM models using the extended multimodal dataset identified similar features and additional MoCA subtests (i. e. MoCA memory and cube subscores) suggesting that poor performance in these cognitive functions increase the risk for conversion to MCI within 3 years. Importantly, these results are also in agreement with evidence from non-ML studies adding to the generalizability of our findings. In this regard, studies using detailed neuropsychological assessments to characterize the profile and sequence of specific cognitive functions in the progression to cognitive impairment in PD previous studies have shown that posterior cortical located visuospatial abilities and frontostriatal represented executive functions are predictors for conversion to MCI and dementia in PD (59–61).

Imaging features ranked as important included the superior cerebellar peduncles (SCPs), cingulate cortex, hippocampus, total brain volume and T1 hypointensities. The SCPs mainly encompass efferent projections of cerebello-thalamo-striato-cortical loops (62). Cerebellar grey matter atrophy and metabolic alterations of the cerebellum have been described in PD (63), even in early stages (64), and have also been associated with cognitive impairment (65, 66), specifically related to executive function (67), dual-task performance (68) as well as verbal and semantic fluency (69). Furthermore, the SCPs receive afferent projections from the noradrenergic locus coeruleus (70, 71), which is centrally implicated in cognitive impairment of attention, working memory and cognitive flexibility in PD (72). While the association of pathological changes in the hippocampus with cognitive impairment is well-established (73), the cingulate cortex has been shown to be a central hub for the integration of motor and cognitive information related to executive function and dual-task performance (74). Age-related increase of T1 hypointensities – analogous to White Matter Hyperintensities (WMHs) – are consistently associated with a decline in cognitive functions, particularly executive functions, attention, and processing speed, even in cognitively unimpaired older adults (75). Finally, decreases in total brain volume have also been shown to be associated with accelerated decline in cognitive functions (76, 77).

Taken together, these imaging-based findings encompass brain regions that are associated with specific cognitive functions (e. g. executive functions, visuospatial abilities, attention, working memory and processing speed), which are well-established to be impaired early in PD and to decline with PD progression (59–61).

For the motor endpoint PI, feature importance ranking using the minimal dataset yielded the same features as for the cognitive endpoint MCI, with higher age and lower MoCA scores predicting conversion to PI. Furthermore, the EBM models using the extended dataset also revealed more detailed information on the features contributing to progression to PI, specifically MDS-UPDRS III subitems (“posture”, “arising from chair” and “rigidity of the right lower extremity”) as well as height and the MoCA sentence repetition subscore. These findings are consistent with a number of previous studies indicating that onset of postural instability is mainly associated with higher age (78–81) and that PD patients with postural instability exhibit worse cognition (78). Furthermore, associations of different MDS-UPDRS III subitems or subscores with axial impairment and postural instability in PD have also been shown in previous studies (82, 83), in particular including gait, posture and balance subitems comparable to the features resulting from our ML-based analysis.

Imaging features resulting from the EBM model for prediction of conversion to PI included volumes of the left hippocampus, bilateral cingulate gyri, left occipital lobe, third ventricle and brain total volume. Besides the well-known role of the hippocampus in memory consolidation (84), it has been shown that the hippocampus is also involved in postural control in aging (85) and that memory impairment is associated with more severe postural instability in PD (86). Similarly, the cingulate cortex has also been shown to be involved in the processing of information related to postural control, e. g. in terms of recognition of unstable postures (87), and notably decreased structural connectivity between fronto-parietal regions including the cingulate gyrus has been shown in the Postural Instability/Gait Difficulty (PIGD) subtype of PD (88). Visuospatial functions – topographically related to occipital regions – have been shown to be impaired in PD patients of the postural instability and gait difficulty type (PIGD) (89). Brain total and third ventricle volumes might be relevant for postural control in a more general context as postural instability has been shown to be an independent marker of brain atrophy (90).

In summary, our findings on the one hand validate that baseline premorbid cognitive and demographic features are relevant for the prediction of progression to the cognitive endpoint MCI, while on the other hand motor, cognitive and demographic features showed relevant contribution for the prediction of progression to the motor endpoint PI. The relevance of this close interaction of motor and cognitive factors and their dual contribution to increased risk of dementia has been recognized in recent years, e.g. in the context of concepts like the “Motor Cognitive Risk syndrome” (MCR) (91–93) or resilience models like the concepts of cognitive and motor reserve (94), which have been shown to be associated with a more rapid progression to postural instability and cognitive impairment in PD (95).

We acknowledge several limitations of our study. First, the data from our local TuePaC cohort were analysed retrospectively. However, TuePaC participants were consecutively recruited closely resembling real world routine clinical care settings. Furthermore, data of the PPMI cohort were collected prospectively and has been widely used for the purpose of validation. Additionally, we did not adjust for systematic differences between the two datasets, which provides a realistic estimate of the model’s performance upon deployment, but may also lead to some loss in performance. Additionally, some information has been lost by harmonizing the two datasets. Moreover, working with real-world clinical data also imposes a challenge in terms of data quality as extendend multidmodal datasets are often not readily available and not easy to be obtained resulting in higher percentages of missing values. However, this is also a realistic reflection of the data quality occurring in divers clinical care settings. We propose that a predictive model necessarily needs to be able to be robust towards different levels of data quality to be useful in routine care. Finally, data for validation of our findings in a closely matched, prospective cohort of early PD patients (LOC Early PD study cohort) will be available shortly.

In conclusion, our study shows that ML models enable predictions at the individual level and facilitate risk assessment for adverse events, which could support personalized clinical decision-making. Thereby, our study exemplifies implications for the implementation of ML methods like EBMs in real world settings of the routine clinical care of PD patients. ML-based predictions using clinico-demographic datasets derived from routine clinical care might allow for a general evaluation of PD progression. In contrast, extended multimodal datasets including imaging features enable a more in-depth phenotyping, allowing a more precise and individualized understanding of factors contributing to PD progression. These advances might support more holistic evaluation and monitoring of PD disease progression facilitating the design of integrated care concepts that promise to address motor and cognitive symptoms of PD patients with more individualized approaches. However, since data used here primarily originated from clinical studies, further validation of these approaches using real-world data particularly from primary care providers is essential. Given current digitalisation efforts, e.g. with the European health data space and the German electronic patient record, access to standardized data from primary care providers that could be used for predicting progression in PD patients comes within reach.

## Methods

### Study design and participants

This study included longitudinal data of PD patients from two independent cohorts:

1. The TuePaC cohort comprises a large dataset of a pooled local cohort of PD patients consecutively recruited between October 2000 and March 2023 at the Department of Neurodegenerative Diseases of the University Hospital Tübingen. The study was approved by the Ethics Committee of the University of Tübingen. All participants gave written informed consent. Study data were collected and managed using REDCap electronic data capture tools hosted at University of Tübingen (96).
2. The PPMI study (www.ppmi-info.org) is an observational, multi-center natural history study of PD leveraging longitudinal demographic, clinical, imaging and digital sensor data as well as biologic and genetic biomarkers since 2010. The study was registered at clinicaltrials.gov (NCT01141023). All participating PPMI sites obtained approval from local ethical review boards as well as written informed consent from all participants prior to inclusion into the study. Data used for the present study was downloaded in January 2023 from the PPMI database (https://ida.loni.usc.edu/). Inclusion criteria for the PPMI study are available on the PPMI website (https://www.ppmi-info.org/study-design/research-documents-and-sops/). It was ensured that these matched the inclusion criteria of the TuePaC cohort (see detailed information on data harmonization in the supplementary methods).

For both cohorts, PD patients with the following criteria were included:

(i) maximum disease duration of 3 years after PD diagnosis, and
(ii) availability of data for the baseline and 3-year follow-up visit (M36).

All patients with a diagnosis of dementia at baseline were excluded from this study.

Furthermore, from the PPMI study cohort we selected a nested sample of participants with available 3D T1 MRI data for additional analyses.

### Multimodal data and feature selection

Two datasets with harmonized features were defined based on overlaps of variables between the TuePaC and PPMI cohort:

1) The Minimal Data Set comprised: baseline age, gender, family history of PD, Hoehn & Yahr stage, Movement Disorder Society-sponsored revision of the Unified Parkinson’s Disease Rating Scale (MDS-UPDRS) part III (97), PD-related motor complications (MDS-UPDRS IV), 15-item geriatric depression scale (GDS) (98), Montréal Cognitive Assessment (MoCA) (99), which are commonly assessed in the routine clinical care of PD patients (see supplementary table 1). Since the MoCA was not available until 2009, MMSE scores for visits without available MoCA were converted into a MoCA equivalent score based on established conversion algorithms (100).
2) The extended dataset contained additional features for PD including medication as well as in-depth motor, clinical, cognitive and laboratory data (supplementary table 1).

The model performances were assessed by comparing (i) the minimal dataset with the extended dataset and (ii) the minimal datasets with and without imaging features respectively.

### Definition of clinical endpoints

Two clinical disease milestones of PD were defined as endpoints for our analysis:

1) Mild Cognitive Impairment (MCI) was defined according to level 1 criteria for MCI by the Movement Disorders Society (MDS) (101) with a score of ≤ 26 points in the MoCA (102). This liberal cut-off was used in order to be most sensitive for early cognitive change, expected to be subtle in the early stage of PD.
2) Postural Instability (PI) was assessed by subitem 12 of the MDS-UPDRS III (score ≥ 1) as a clinical motor endpoint.

### Brain volumetry

For the PPMI imaging subcohort, 3D T1-weighted Magnetization Prepared Rapid Acquisition with Gradient Echoes (MPRAGE) data was downloaded from the IDA LONI platform (PD cases at baseline n=492) and processed using a recently established automated volumetry pipeline (47) generating 34 volumetric features.

For detailed information on the PPMI MRI protocol and acquisition parameters we refer to the online documentation on the PPMI website (https://www.ppmi-info.org/study-design/research-documents-and-sops/).

### Data pre-processing and definition of feature list

Data was excluded based on the following criteria: (1) Features in the study cohort pooled from the two cohorts with more than 40% missing baseline data (see supplementary table 1 for details), (2) patients with more than 50% missing baseline data (3) patients with missing information on the respective endpoints at M36 (TuePaC: MCI n=37.1%, PI n=50.9%; PPMI: MCI n=3.7%, PI n=57%). Categorical data underwent one-hot encoding (OHE). Outliers were removed using Interquartile Range (1.5 * IQR) (to exclude most likely measurement errors). To reduce the number of missing data points ethnicity was set as “Caucasian” and family history as “negative” in the TuePaC cohort (see supplementary table 1 for details).

#### Imputation

After selection of features and patients for the final dataset K-Nearest Neighbour (KNN), Mean/Mode imputation and MICE/Missforest imputation (MICE: https://www.jstatsoft.org/article/view/v045i03) were compared by calculating the Normalised Root Mean Squared Error (NRMSE), ‘MAE’ (Mean Absolute Error), the ‘R-squared score’, the ‘Mean Squared Logarithmic Error’ and the ‘Explained Variance score’ for 100 iterations with different repeated imputations (see Supplementary table 2). MICE and Missforest imputation were finally selected for imputation due to their ability to handle complex data (see Supplementary figures 1-3). Hyperparameters were tuned using RandomSearchCV with n_iter=100 on the generated dataset.

#### Feature Engineering

Feature engineering procedures, including feature binning and mean target encoding, were applied to enhance the datasets’ informational value. Additional transformations, such as encoding family anamnesis and ethnicity, were implemented to reduce feature dimensionality and increase information density.

### Explainable Boosting Machine Learning

#### Training & used algorithms

Python (version 3.8.10) was used in this study for constructing the analysis pipeline and tuning the hyperparameters using a range of open-source libraries such as NumPy, pandas, scikit-learn, imbalanced-learn, interpret, scipy, seaborn, and matplotlib. Figures were generated using R (version 4.4.0) and open-source libraries tidyverse, patchwork and ggpubr. Models were trained based on PPMI as training and TuePaC as independent test data set, while a random split (60% training and 40% test data) was applied for the imaging sub-cohort analysis. To cope with overfitting 5-fold cross-validation was used.

EBMs were selected as the primary model for their unique ability to balance predictive performance with interpretability, unlike traditional models that often require a trade-off between these aspects (58, 103). EBMs discretize continuous variables, encode categorical variables, and assign score functions to each feature, which are summed with an intercept to produce predictions. This additive strucutre ensures transparency and facilitates model debugging (104, 105).

#### Testing

The different models were applied for predicting classes (MCI or PI conversion) on the test dataset, resulting in various evaluation metrics (AUC, Accuracy, Sensitivity, Specificity), as well as additional evaluation elements like confusion matrices.

#### Resampling methods

Unequal class distributions can bias ML models machine learning models toward the majority class. To mitigate this in our dataset, we employed oversampling and class weighting. Oversampling increases minority class representation, while class weighting makes the model more sensitive to minority class errors. For oversampling, we tested the RandomOverSampler, SMOTE, and several variants of it (BorderlineSMOTE, KMeansSMOTE, SVMSMOTE), as well as ADASYN, all from the imbalanced-learn package. Detailed descriptions of these methods are provided in the supplementary material (see Supplementary Methods). The optimal performance was achieved using the ADASYN algorithm with k_neighbors=6, in conjunction with a class weight of 10:1.

#### Feature Selection

Several feature selection algorithms (DropConstantFeatures, DropDuplicateFeatures, SelectByTargetMeanPerformance, SelectBySingleFeaturePerformance, SmartCorrelatedSelection, DropCorrelatedFeatures, SelectByShuffling, RecursiveFeatureAddition, RecursiveFeatureElimination (RFE) and RFE with cross-validation (RFECV)) were tested, but didn’t provide significant benefits in model performance. Therefore, only features with a Pearson correlation greater than 80% were dropped from the model. In such cases, features with more missing values were dropped, whereas correlated features with fewer missing values were kept for the analysis.

### Statistical analysis

Statistical analysis was performed using Python 3.8.10 and scipy 1.5.4 software. Mann-Whitney-U test was used for continuous variables and χ^2^ test was used for categorical data. All statistical tests were two-sided and *p* values ≤ 0.05 were considered statistically significant.

## Data availability

Data of this study including data from the TuePaC cohort are available upon request to the corresponding author. Data from the PPMI study used in the present study were downloaded on January 27, 2023 from the PPMI database (www.ppmi-info.org/data).

## Code availability

The code programmed for and used in this study is accessible in the following GitHub repository: https://github.com/meDIC-UKT/LOC-PD.

## Acknowledgements

This work is part of the DIFUTURE (Data Integration for Future Medicine) project funded by the German Ministry of Education and Research (Bundesministerium für Bildung und Forschung, BMBF), Grant 01ZZ1804[A-I].

Data used in the preparation of this article was obtained on 2023-01-27 from the Parkinson’s Progression Markers Initiative (PPMI) database (www.ppmi-info.org/access-data-specimens/download-data), RRID:SCR_006431. For up-to-date information on the study, visit www.ppmi-info.org.

PPMI – a public-private partnership – is funded by the Michael J. Fox Foundation for Parkinson’s Research, and funding partners; including including 4D Pharma, Abbvie, AcureX, Allergan, Amathus Therapeutics, Aligning Science Across Parkinson’s, AskBio, Avid Radiopharmaceuticals, BIAL, BioArctic, Biogen, Biohaven, BioLegend, BlueRock Therapeutics, Bristol-Myers Squibb, Calico Labs, Capsida Biotherapeutics, Celgene, Cerevel Therapeutics, Coave Therapeutics, DaCapo Brainscience, Denali, Edmond J. Safra Foundation, Eli Lilly, Gain Therapeutics, GE HealthCare, Genentech, GSK, Golub Capital, Handl Therapeutics, Insitro, Jazz Pharmaceuticals, Johnson & Johnson Innovative Medicine, Lundbeck, Merck, Meso Scale Discovery, Mission Therapeutics, Neurocrine Biosciences, Neuron23, Neuropore, Pfizer, Piramal, Prevail Therapeutics, Roche, Sanofi, Servier, Sun Pharma Advanced Research Company, Takeda, Teva, UCB, Vanqua Bio, Verily, Voyager Therapeutics, the Weston Family Foundation and Yumanity Therapeutics.

The AIRAmed GmbH provided processing of imaging data as part of a research agreeement free of charge without any influence on design, data analysis or interpretation of the present study.

## Contributions

B.R. and I.L.-S. defined the study aims and objectives. B.S., B.R., C.K., K.B., S.B. and I.L-S. contributed to study conceptualization and coordination. B.S. designed and primarily programmed the ML pipeline. C.K. and L.A. contributed to the ML pipeline programming and analyses. B.S., B.R. and I.L.-S. prepared the draft of the manuscript, which was critically reviewed and approved by all authors. B.S., C.K., L.A. and S.B prepared parts of the methods and results section of the manuscript. B.R., C.K., R.W., I.W., S.L., C.S., C.D., C.E., T.G. and K.B. contributed to data collection and/or curation. B.S., B.R., C.K., L.A., S.B. and I.L.-S. analyzed the data and interpreted the results. B.B. and T.L. processed the imaging data. S.B. and I.L.-S. supervised the study.

B.S., B.R. and C.K. contributed equally and share first authorship. S.B. and I.L.-S. contributed equally as senior authors.

## Abbreviations

MDS-UPDRS: Movement Disorders society sponsored revision of the Unified Parkinson’s Disease Rating Scale
MoCA: Montréal Cognitive Assessment
MCI: Mild Cognitive Impairment
PI: Postural Instability
ML: Machine Learning
EBM: Explaniable Boosting Machine

## DIFUTURE consortium

Prof. Dr. Martin Boeker ^1^, Prof. Dr. Ulrich Mansmann ^2^, Prof. Dr. Oliver Kohlbacher ^3, 4, 5^, Prof. Dr. F. Kramer ^6^, Prof. Dr. Hans Kestler ^7^

^1^AI and Informatics in Medicine, TUM University Hospital (MRI), Technical University of Munich (TUM), Munich, Germany.

^2^Departement of Medical Information Processing, Biometry, and Epidemiology, University Hospital Munich (KUM), Ludwig Maximilian University of Munich (LMU), Munich, Germany.

^3^Applied Bioinformatics, Dept. of Computer Science, University of Tübingen (EKUT), Tübingen, Germany

^4^Institute for Bioinformatics and Medical Informatics, University of Tübingen (EKUT), Tübingen, Germany

^5^Translational Bioinformatics, University Hospital Tübingen (UKT), Tübingen, Germany

^6^IT Infrastructure for Translational Medical Research, University Hospital Augsburg (UKA), University of Augsburg (UA), Augsburg, Germany

^7^Institute for Medical Systems Biology, Ulm University Hospital (UKU), Ulm University, Ulm, Germany.

