## Supplementary Material for "Explainable machine learning identifies predictors for postural instability and cognitive impairment in early Parkinson’s Disease"

### **Supplementary methods**

#### **Data set**

A detailed description of the data and the features used for each feature set (minimal, extended and imaging) is shown in Supplementary Table 1. Additionally, information about why certain features have been included/excluded can be found in this table.

Since the MoCA only became available in 2009, Mini–Mental Status Examination (MMSE) scores have been converted to MoCA-equivalent scores using a previously established conversion algorithm (1) if necessary.

The following demographic data, clinical scores and endpoints were selected as features for the machine learning analysis.

*Demographics.* Age, gender, ethnicity, disease duration, and family history for PD were assessed based on predefined structured questionnaires/CRFs.

*Motor symptoms & complications.* Severity of motor symptoms was assessed using the Unified Parkinson’s Disease Rating Scale part III (UPDRS III) from 2003 to 2008 and the Movement Disorder Society (MDS)–UPDRS part III from 2009 on. Presence or occurrence of postural instability was assessed by subitem 12 of the MDS-UPDRS III as a clinical motor endpoint at year 3. The UPDRS part IV or the MDS-UPDRS part IV were used to assess the severity of motor complications of PD including subitem scores for freezing of gait, dyskinesias and Off dystonia symptoms.

The Levodopa-equivalent dose (LEDD) was calculated according to established conversion factors for levodopa-equivalent doses (2).

*Cognition.* Cognitive function was assessed by the Mini–Mental Status Examination (MMSE) and/or the Montréal Cognitive Assessment (MoCA).

*Depression.* The Geriatric Depression Scale (GDS) was used to assess depressive symptoms.

#### **Data Harmonization and Visit Structure Alignment**

Due to differences in data structures, variable nomenclature, and other discrepancies between the two data sources, both cohorts underwent a harmonization process prior to analysis. This process was performed using a custom mapping script specifically developed for this study.

To achieve comparability, a standardized visit schedule typical of longitudinal studies—comprising one baseline visit and up to three annual follow-up visits—was defined as the target framework. Since the TuePaC data were derived from routine clinical practice, data points were retrospectively grouped into artificial visits corresponding to this schedule, which was already inherently present in the PPMI dataset. The entire observation period following each patient’s initial clinical presentation was considered.

For the baseline visit, all measurements collected within the first three months after initial presentation were included. For follow-up visits at Month 12 (M12), Month 24 (M24), and Month 36 (M36), a ±6-month window around the theoretical visit date was applied. For example, data recorded 6–17 months after the initial presentation were assigned to the artificial M12 visit. In cases where multiple measurements were available for the same variable within a visit window due to patients presenting on multiple occasions the most recent measurement within that interval was retained.

Medication data were handled differently for the two cohorts. In the Tuebingen dataset, medications were recorded per visit, including active ingredient and dosage. In contrast, the PPMI dataset provided more comprehensive information, with medication start and end dates recorded. For harmonization, current medication status at each visit in the PPMI cohort was derived from these time-stamped medication histories.

#### **Data pre-processing**

##### ***Missing Data Assessment***

All variables were systematically assessed for the proportion and absolute number of missing values. Features with more than 40% missing baseline data across the pooled study cohort (see Supplementary Table 1 for details) were excluded from further analyses. Variables that were rarely recorded were removed, whereas variables typically expected to be available, such as age or sex, were retained despite some missingness.

Patient-level exclusions were performed as follows:

1. Patients with more than 50% missing baseline data were removed.

2. Patients lacking primary endpoint data at Month 36 were excluded (TuePaC: Montreal Cognitive Assessment [MoCA] missing in 37.1% of cases; Unified Parkinson’s Disease Rating Scale [UPDRS] missing in 50.9%; PPMI: MoCA missing in 3.7%, UPDRS missing in 57.0%). Records missing these endpoints were not used for model development.

To reduce baseline missingness in specific variables, predefined imputations were applied in the TuePaC dataset: missing ethnicity values were set to “Caucasian,” and missing family history entries were set to “negative.”

Categorical variables were transformed using one hot encoding. Outliers, considered likely to represent measurement errors, were identified and removed using the interquartile range method (threshold: 1.5 × IQR from the first and third quartiles).

##### ***Imputation***

Owing to the fact that we used clinical routine data which in general is characterized by reduced data quality and higher number of missingness than clinical study data, we underwent a thorough imputation evaluation, comparing different imputation methods. From the original dataset (retro+PPMI combined), a dataset without missing data was created by removing all records with missing data. The same amount of data from each feature as in the original complete dataset was removed to create a copy with missing data. Three different imputation methods were applied to this dataset (Supplementary Figure 2), namely K-Nearest-Neighbor (KNN), mean/mode imputation and MICE/Missforest imputation (3), compared with each other based on their Normalised Root Mean Squared Error (NRMSE). In addition, the ‘MAE’ (Mean Absolute Error), the ‘R-squared score’, the ‘Mean Squared Logarithmic Error’ and the ‘Explained Variance score’ were calculated (Supplementary Table 2).

Several estimators of the iterative imputer were tested, and their NRMSEs were compared. MICE, MissForest, Decision Tree, K-Nearest Neighbour (KNN) and Linear Regression. The imputation process was repeated 100 times (see Supplementary Figure 3).

Hyperparameters of MICE and Missforest imputation were tuned using RandomSearchCV with n_iter=100 on the generated dataset.

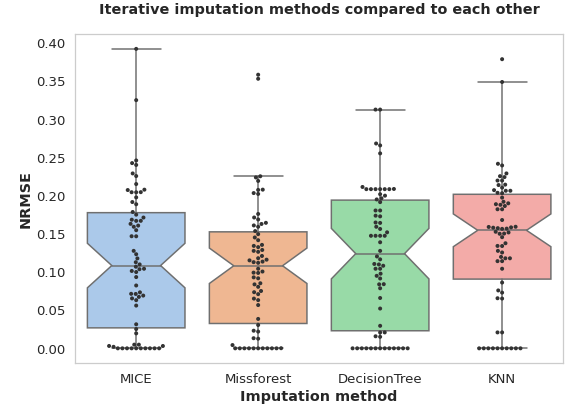

**Supplementary Figure 1.** **Comparison of iterative imputation methods** based on Normalized Root Mean Squared Error (NRMSE) across repeated imputations. MICE and MissForest show the lowest NRMSE across the imputation methods compared.

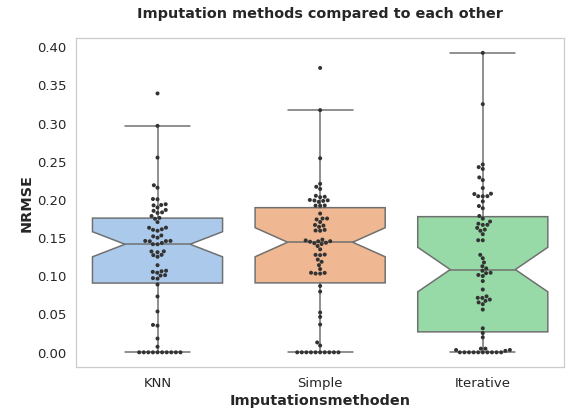

**Supplementary Figure 2.** **Comparison of KNN, Simple, and Iterative imputers** based on Normalized Root Mean Squared Error (NRMSE) applied to datasets with original and synthetically removed missing values. Iterative imputer shows the lowest NRMSE, whereas KNN and a simple mean/mode imputation perform slightly worse.

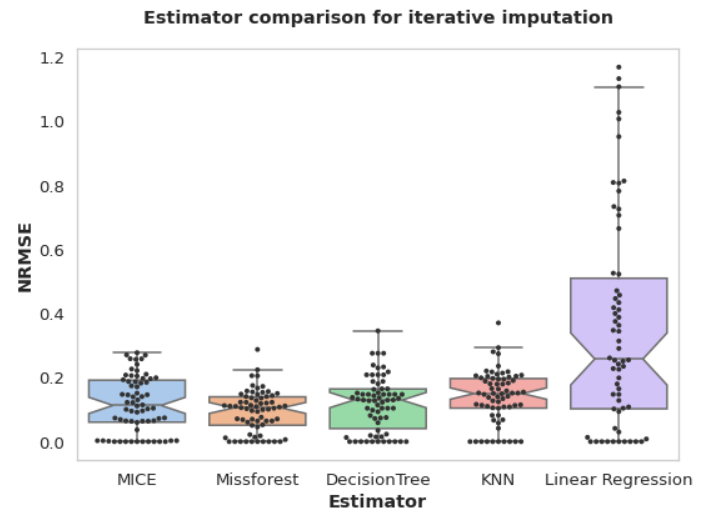

**Supplementary Figure 3. Comparison of imputation estimators** used for iterative imputation. MissForest and MICE performed similarly and achieved the best results, while DecisionTree and KNN showed slightly lower performance. Linear Regression yielded the poorest imputation.

##### ***Feature engineering***

Feature engineering can significantly impact model performance by creating new features and/or encoding existing features in a different way. We utilized several techniques to the data used.

*Age* and *MoCA* score were binned into categories (Age: 18-30, 30-50, 50-70, 70+, MoCA score: <10, 11-17, 18-25, >25) and ordinally encoded afterwards.

We also created derived features: A binary *retiremen*t feature was created, categorizing patients as retired if their age exceeded 65 years. This age threshold was chosen based on common retirement practices in the study regions. To capture the complexity of patient medication regimens, we calculated a *medication count* feature, representing the total number of medications prescribed during each visit. Given the ethnically homogeneous nature of the TuePaC dataset, we simplified the ethnicity variable into a binary *Caucasian* feature (yes/no).

To quantify the genetic risk associated with *family history* of Parkinson's disease, we developed an ordinal encoding scheme based on the degree of relatedness to affected family members:

- 0: No family history of Parkinson's disease
- 1: First-degree relatives (parents, siblings, or children) diagnosed with Parkinson's disease
- 2: Second-degree relatives (grandparents) with Parkinson's disease
- 3: Third-degree relatives (uncles or aunts) affected by Parkinson's disease

Target encoding was applied to boolean, ordinal, and categorical features. In this method, the original feature values are replaced by the mean of the target variable corresponding to each unique feature value.

For numerical features, temporal changes were quantified by calculating the difference between values recorded at a given visit and those from the previous visit. However, these derived temporal features were not analyzed in detail, as we focused on predictions based on baseline data.

#### **Machine Learning**

##### ***Model Implementation and Software Specifications***

Python (version 3.8.10) was used in this study for constructing the analysis pipeline and tuning the hyperparameters using a range of open-source libraries such as NumPy, pandas, scikit-learn, imbalanced-learn, interpret, seaborn, and matplotlib. Figures were generated using R (version 4.4.0) and  open-source libraries tidyverse, patchwork and ggpubr. Models were trained based on PPMI as training and retro Tübingen DS as independent test data set, or, as in the case of the imaging sub-cohort analysis, a random split with 60% training and 40% test data was being applied. To cope with overfitting 5-fold cross-validation was used.

In this study, Explainable Boosting Machines (EBMs) were selected as the primary modeling approach due to their ability to combine strong predictive performance with full interpretability, avoiding the common trade-off between simple but transparent models (e.g., logistic regression) and complex “black-box” methods (e.g., gradient-boosted trees, neural networks). Built on the Generalized Additive Models plus interactions (GA²M) framework, EBMs learn separate shape functions for individual features, capturing nonlinearities by discretizing continuous variables into bins, and can optionally incorporate pairwise interaction terms. Predictions are generated as the sum of these learned feature contributions plus a model intercept, allowing direct visual inspection of each feature’s influence and simplifying the debugging and validation process.

For this analysis, EBMs were trained and tuned using a randomized search over a pre-defined hyperparameter space. The following parameters were optimized:

- **outer_bags**: uniformly sampled integers between 1 and 50;
- **inner_bags**: uniformly sampled integers between 1 and 50;
- **max_bins** (number of discrete bins for continuous features): uniformly sampled integers between 2 and 500;
- **early_stopping_rounds**: uniformly sampled integers between 10 and 100;
- **early_stopping_criteria**: grid search across values from 2,500 to 7,500 in steps of 500;
- **max_leaves** (maximum number of leaves per tree in each boosting iteration): values between 2 and 5.

Randomized search was performed with cross-validation to identify the hyperparameter combination yielding optimal model performance on the training set, while guarding against overfitting. This configuration ensured a balance between model complexity and generalizability, while preserving the interpretive clarity inherent to EBMs.

##### ***Testing***

Various models were applied to predict MCI or PI conversion and evaluated on the independent test dataset, resulting in various evaluation metrics (AUC, Accuracy, Sensitivity, Specificity), as well as additional evaluation elements (like confusion matrices, Receiver Operator Characteristics (ROC)-AUC curves and corresponding plots. The Youden index (Sensitivity + Specificity - 1) was used for selecting the best performing models.

#### ***Resampling methods***

Data sets with unequal class distributions can lead to problems in various machine learning models, in that the minority class is ignored and the trained model focuses only on the majority class. Within the last decades, several solutions to this problem have been established, namely undersampling, oversampling, and class weighting. In undersampling, samples of the majority class are randomly removed from the dataset, while in oversampling synthetic samples of the minority class are generated. In class weighting, the minority class is assigned a higher weight, i.e., misclassifications by the model are penalized more severely with a higher loss. Due to the small dataset size by ML standards, we only considered methods of oversampling, as well as different class weighting.

For oversampling, we tested the RandomOverSampler, SMOTE, and several variants of it (BorderlineSMOTE, KMeansSMOTE, SVMSMOTE), as well as ADASYN all from the imbalanced-learn package:

- RandomOverSampler: picks samples of the minority class at random with replacement
- SMOTE : uses k-nearest neighbors to create synthetic examples of the minority class by extrapolating between existing examples.
- BorderlineSMOTE : only minority samples near the decision boundary are oversampled
- KMeansSMOTE : clusters samples and selects clusters for oversampling
- SVMSMOTE : uses an SVM algorithm to pick samples for oversampling
- ADASYN : uses a weighted distribution which weights samples higher that are harder to classify.

We obtained the best results with ADASYN with k_neighbors=6, as well as using different class weights.

##### ***Feature selection***

Several feature selection algorithms (Recursive Feature Elimination/Addition, SelectByTargetMeanPerformance, ...) from the feature-engine package were tested:

- DropConstantFeatures: drops constant features from the data
- DropDuplicateFeatures: drops duplicate features from the data
- SelectByTargetMeanPerformance: replaces feature values by the mean value of the target variable per category and selects features based on the performance of this new feature.
- SelectBySingleFeaturePerformance: trains a ML model for each single feature and evaluates the performance for this feature.
- SmartCorrelatedSelection: finds correlated features and drops them in a smart way (e.g. by keeping the features with the least missing values)
- DropCorrelatedFeatures: finds correlated features and drops them.
- RFECV (Recursive feature elimination with cross-validation): Trains an estimator recursively and prunes the least important features. Process is repeated until the stop criterion is reached.
- Recursive Feature Addition: Similar to recursive feature elimination but adds features iteratively instead of dropping them based on their performance.

Feature selection didn't provide significant benefits in model performance. Therefore, only features with a Pearson correlation greater than 80% were dropped from the model.

### **Supplementary results**

#### **Performance using other Machine Learning models**

In addition to the primary model, we trained alternative machine learning algorithms, comprising logistic regression, random forest, Explainable Boosting Machine (EBM), multilayer perceptron (MLP), and XGBoost, on datasets collected at baseline (M0) and at 12-, 24-, and 36-month (M12, M24, M36) follow-up visits. Classification results for mild cognitive impairment (MCI) and postural instability (PI) are presented in Supplementary Tables 3a and 3b, respectively.

#### **EBM model runs**

Details of the Explainable Boosting Machine (EBM) runs employed for the primary model, along with the corresponding analysis-specific hyperparameters, are provided in Supplementary Tables 4a (MCI) and 4b (PI). Additionally, results pertaining to the imaging cohort, using both available structured imaging data and clinical data only are reported in Supplementary Tables 5a and 5b.

Split size refers to the proportion of data allocated to the training and testing subsets, while sample weights specify the relative importance assigned to positive and negative classes.

| **Features** | **Mild Cognitive Impairment (MCI)** | | | | | | **Postural Instability (PI)** | | | | | |
| --- | --- | --- | --- | --- | --- | --- | --- | --- | --- | --- | --- | --- |
|  | Minimal | | | Extended | | | Minimal | | | Extended | | |
|  | Clinical cohort | Imaging  Cohort | | Clinical cohort | Imaging  Cohort | | Clinical cohort | Imaging  Cohort | | Clinical cohort | Imaging  Cohort | |
|  |  | w/ imaging | w/o imaging |  | w/  imaging | w/o  imaging |  | w/  imaging | w/o imaging |  | w/  imaging | w/o imaging |
| N of PD cases analysed | 539 | 100 | 113 | 400 | 113 | 113 | 558 | 87 | 101 | 427 | 101 | 101 |
| N of features included into the analysis | 4 | 17 | 7 | 69 | 65 | 56 | 4 | 24 | 7 | 56 | 54 | 45 |
| **Demographic and individual characteristics** | | | | | | | | | | | | |
| *Age* | + | + | + | + | + | + | + | + | + | + | + | + |
| *Gender* | + | + | + | + | + | + | + | + | + | + | + | + |
| Employed |  |  |  | - MV=59% | - MV=87% | - MV=87% |  |  |  | - MV=61% | - MV=87% | - MV=87% |
| Ethnicity |  |  |  | - r>0.80 | - MV=74% | - r>0.80 |  |  |  | - r>0.80 | - MV=74% | - MV=74% |
| Height |  |  |  | + | + | + |  |  |  | + | + | + |
| Weight |  |  |  | + | + | + |  |  |  | + | + | + |
| *Family history* | + | - r>0.80 | + | + | + | + | + | - r>0.80 | + | + | + | + |
| **Motor function** | | | | | | | | | | | | |
| *mod. Hoehn & Yahr* | - MV=53% | + | + | + | + | + | - MV=53% | + | + | + | + | + |
| UPDRS (Total score) I |  |  |  | - MV=51% | + | + |  |  |  | - MV=54% | + | + |
| UPDRS (Total score) II |  |  |  | + | + | + |  |  |  | + | + | + |
| *UPDRS (Total score) III* | - MV=52% | + | + | - r>0.80 | - r>0.80 | - r>0.80 | - MV=42% | + | + | - MV=42% | - r>0.80 | - r>0.80 |
| 3.1 Speech |  |  |  | + | + | - r>0.80 |  |  |  | + | + | - r>0.80 |
| 3.2 Facial Expression |  |  |  | + | + | + |  |  |  | + | + | + |
| 3.3 Rigidity Neck |  |  |  | + | + | + |  |  |  | + | + | + |
| 3.3 Rigidity RUE |  |  |  | + | + | + |  |  |  | - r>0.80 | - r>0.80 | + |
| 3.3 Rigidity LUE |  |  |  | - r>0.80 | + | - r>0.80 |  |  |  | + | - r>0.80 | + |
| 3.3 Rigidity RLE |  |  |  | + | + | + |  |  |  | + | + | + |
| 3.3 Rigidity LLE |  |  |  | + | - r>0.80 | - r>0.80 |  |  |  | + | + | - r>0.80 |
| 3.4 Finger Tapping R |  |  |  | + | + | + |  |  |  | + | + | + |
| 3.4 Finger Tapping L |  |  |  | + | + | + |  |  |  | + | + | + |
| 3.5 Hand Movements R |  |  |  | - r>0.80 | - r>0.80 | + |  |  |  | + | + | - r>0.80 |
| 3.5 Hand Movements L |  |  |  | + | + | + |  |  |  | + | + | - r>0.80 |
| 3.6 Pronation-Supination Movements of Hands R |  |  |  | + | + | + |  |  |  | + | + | + |
| 3.6 Pronation-Supination Movements of Hands L |  |  |  | + | + | + |  |  |  | + | + | - r>0.80 |
| 3.7 Toe Tapping R |  |  |  | - r>0.80 | - r>0.80 | - r>0.80 |  |  |  | + | + | + |
| 3.7 Toe Tapping L |  |  |  | + | + | + |  |  |  | - r>0.80 | + | + |
| 3.8 Leg Agility R |  |  |  | + | + | + |  |  |  | + | - r>0.80 | + |
| 3.8 Leg Agility L |  |  |  | + | + | + |  |  |  | + | + | + |
| 3.9 Arising from Chair |  |  |  | + | + | + |  |  |  | - r>0.80 | + | - r>0.80 |
| 3.10 Gait |  |  |  | - r>0.80 | - r>0.80 | - r>0.80 |  |  |  | - r>0.80 | - r>0.80 | - r>0.80 |
| 3.11 Freezing of Gait |  |  |  | + | + | + |  |  |  | + | + | + |
| 3.12 Postural Stability |  |  |  | + | + | + |  |  |  | + | + | + |
| 3.13 Posture |  |  |  | - r>0.80 | - r>0.80 | - r>0.80 |  |  |  | - r>0.80 | - r>0.80 | - r>0.80 |
| 3.14 Body Bradykinesia |  |  |  | - r>0.80 | + | + |  |  |  | + | + | + |
| 3.15 Postural Tremor of Hands R |  |  |  | + | + | + |  |  |  | + | + | + |
| 3.15 Postural Tremor of Hands L |  |  |  | + | + | - r>0.80 |  |  |  | + | - r>0.80 | - r>0.80 |
| 3.16 Kinetic Tremor of the Hands R |  |  |  | - r>0.80 | - r>0.80 | - r>0.80 |  |  |  | + | - r>0.80 | + |
| 3.16 Kinetic Tremor of the Hands L |  |  |  | - r>0.80 | - r>0.80 | - r>0.80 |  |  |  | + | - r>0.80 | + |
| 3.17 Rest Tremor Amplitude Neck |  |  |  | + | + | + |  |  |  | + | - r>0.80 | - r>0.80 |
| 3.17 Rest Tremor Amplitude RUE |  |  |  | + | - r>0.80 | + |  |  |  | + | + | + |
| 3.17 Rest Tremor Amplitude LUE |  |  |  | + | - r>0.80 | - r>0.80 |  |  |  | + | - r>0.80 | - r>0.80 |
| 3.17 Rest Tremor Amplitude RLE |  |  |  | + | - r>0.80 | - r>0.80 |  |  |  | + | - r>0.80 | + |
| 3.17 Rest Tremor Amplitude LLE |  |  |  | + | + | + |  |  |  | + | + | + |
| 3.18 Constancy of Rest Tremor |  |  |  | + | + | + |  |  |  | + | + | + |
| **Motor complications** | | | | | | | | | | | | |
| *UPDRS IV (Total score)* | - MV=72% | - MV=54% | - MV=54% | - MV=72% | - MV=54% | - MV=54% | - MV=69% | - MV=54% | - MV=54% | - MV=69 | - MV=54% | - MV=54% |
| UPDRS IV.1 |  |  |  | - MV=69% | - MV=100% | - MV=100% |  |  |  | - MV=67 | - MV=100% | - MV=100% |
| UPDRS IV.2 |  |  |  | - MV=68% | - MV=100% | - MV=100% |  |  |  | - MV=66 | - MV=100% | - MV=100% |
| UPDRS IV.3 |  |  |  | - MV=68% | - MV=100% | - MV=100% |  |  |  | - MV=66 | - MV=100% | - MV=100% |
| UPDRS IV.4 |  |  |  | - MV=68% | - MV=100% | - MV=100% |  |  |  | - MV=66 | - MV=100% | - MV=100% |
| UPDRS IV.5 |  |  |  | - MV=68% | - MV=100% | - MV=100% |  |  |  | - MV=66 | - MV=100% | - MV=100% |
| Dyskinesien choratisch |  |  |  | + | - r>0.80 | - r>0.80 |  |  |  | + | - r>0.80 | - r>0.80 |
| Dystonie |  |  |  | - MV=81% | - MV=99% | - MV=99% |  |  |  | - MV=82 | - MV=99% | - MV=99% |
| motor complications: end-of-dose akinesia / wearing off |  |  |  | - MV=75% | - MV=100% | - MV=100% |  |  |  | - MV=71 | - MV=100% | - MV=100% |
| motor complications: on-freezing |  |  |  | + | - r>0.80 | - r>0.80 |  |  |  | + | - r>0.80 | - r>0.80 |
| **Montreal Cognitive Assessment (MoCA)** | | | | | | | | | | | | |
| *MoCA (Total score)* | + | + | + | + | + | + | + | + | + | + | + | + |
| MoCA trail making |  |  |  | + | + | + |  |  |  | + | - r>0.80 | - r>0.80 |
| MoCA clock contour |  |  |  | + | + | + |  |  |  | + | + | + |
| MoCA clock numbers |  |  |  | + | + | + |  |  |  | + | - r>0.80 | - r>0.80 |
| MoCA clock hands |  |  |  | + | + | + |  |  |  | + | - r>0.80 | - r>0.80 |
| MoCA cube |  |  |  | + | + | + |  |  |  | + | - r>0.80 | - r>0.80 |
| MoCA naming |  |  |  | + | + | + |  |  |  | + | - r>0.80 | + |
| MoCA digit span |  |  |  | + | + | + |  |  |  | + | + | - r>0.80 |
| MoCA letter A tapping |  |  |  | + | + | + |  |  |  | + | + | + |
| MoCA substraction |  |  |  | + | + | + |  |  |  | + | + | + |
| MoCA sentence repetition |  |  |  | + | + | + |  |  |  | + | + | - r>0.80 |
| MoCA letter F fluency |  |  |  | + | + | + |  |  |  | + | - r>0.80 | - r>0.80 |
| MoCA abstraction |  |  |  | + | + | + |  |  |  | + | - r>0.80 | - r>0.80 |
| MoCA memory |  |  |  | + | + | + |  |  |  | + | + | + |
| MoCA orientation |  |  |  | + | + | + |  |  |  | + | - r>0.80 | - r>0.80 |
| **Geriatric Depression Scale (GDS)** | | | | | | | | | | | | |
| *Geriatric Depression Scale (Total score)* | -  MV=54% | + | + | + | + | + | - MV=42 | + | + | - MV=42% | + | + |
| GDS.1 - Are you basically satisfied with your life? |  |  |  | + | - r>0.80 | - r>0.80 |  |  |  | - MV=42% | - r>0.80 | - r>0.80 |
| GDS.2 - Have you dropped many of your activities and interests? |  |  |  | + | - r>0.80 | - r>0.80 |  |  |  | - MV=42% | - r>0.80 | - r>0.80 |
| GDS.3 - Do you feel that your life is empty? |  |  |  | + | - r>0.80 | - r>0.80 |  |  |  | - MV=42% | - r>0.80 | - r>0.80 |
| GDS.4 - Do you often get bored? |  |  |  | + | - r>0.80 | - r>0.80 |  |  |  | - MV=42% | - r>0.80 | - r>0.80 |
| GDS.5 - Are you in good spirits most of the time? |  |  |  | + | - r>0.80 | - r>0.80 |  |  |  | - MV=42% | - r>0.80 | - r>0.80 |
| GDS.6 - Are you afraid that something bad is going to happen to you? |  |  |  | + | - r>0.80 | - r>0.80 |  |  |  | - MV=42% | - r>0.80 | - r>0.80 |
| GDS.7 - Do you feel happy most of the time? |  |  |  | + | - r>0.80 | - r>0.80 |  |  |  | - MV=42% | - r>0.80 | - r>0.80 |
| GDS.8 - Do you often feel helpless? |  |  |  | + | - r>0.80 | - r>0.80 |  |  |  | - MV=42% | - r>0.80 | - r>0.80 |
| GDS.9 - Do you prefer to stay at home, rather than going out and doing new things? |  |  |  | + | - r>0.80 | - r>0.80 |  |  |  | - MV=42% | - r>0.80 | + |
| GDS.10 - Do you feel you have more problems with memory than most? |  |  |  | + | - r>0.80 | - r>0.80 |  |  |  | - MV=42% | - r>0.80 | - r>0.80 |
| GDS.11 - Do you think it is wonderful to be alive now? |  |  |  | + | - r>0.80 | - r>0.80 |  |  |  | - MV=42% | - r>0.80 | - r>0.80 |
| GDS.12 - Do you feel pretty worthless the way you are now? |  |  |  | + | - r>0.80 | - r>0.80 |  |  |  | - MV=42% | + | - r>0.80 |
| GDS.13 - Do you feel full of energy? |  |  |  | + | - r>0.80 | - r>0.80 |  |  |  | - MV=42% | - r>0.80 | - r>0.80 |
| GDS.14 - Do you feel that your situation is hopeless? |  |  |  | + | - r>0.80 | + |  |  |  | - MV=42 | - r>0.80 | + |
| GDS.15 - Do you think that most people are better off than you are? |  |  |  | + | - r>0.80 | - r>0.80 |  |  |  | - MV=42 | - r>0.80 | - r>0.80 |
| **Antiparkinson medication** | | | | | | | | | | | | |
| Amantadin |  |  |  | - MV=98% | - MV=92% | - MV=92% |  |  |  | - MV=98% | - MV=92% | - MV=92% |
| Bromocriptin |  |  |  | - MV=100% | - MV=100% | - MV=100% |  |  |  | - MV=100% | - MV=100% | - MV=100% |
| Budipin |  |  |  | - MV=100% | - MV=100% | - MV=100% |  |  |  | - MV=100% | - MV=100% | - MV=100% |
| Cabergolin |  |  |  | - MV=100% | - MV=100% | - MV=100% |  |  |  | - MV=100% | - MV=100% | - MV=100% |
| Entacapon |  |  |  | - MV=99% | - MV=99% | - MV=99% |  |  |  | - MV=99% | - MV=99% | - MV=99% |
| LEDD |  |  |  | + | + | + |  |  |  | + | + | + |
| Levodopa (direct) |  |  |  | - MV=90% | - MV=76% | - MV=76% |  |  |  | - MV=89% | - MV=76% | - MV=76% |
| Levodopa (retard) |  |  |  | - MV=100% | - MV=100% | - MV=100% |  |  |  | - MV=100% | - MV=100% | - MV=100% |
| Opicapon |  |  |  | - MV=100% | - MV=100% | - MV=100% |  |  |  | - MV=100% | - MV=100% | - MV=100% |
| Pergolid |  |  |  | - MV=100% | - MV=100% | - MV=100% |  |  |  | - MV=100% | - MV=100% | - MV=100% |
| Piribedil |  |  |  | - MV=99% | - MV=100% | - MV=100% |  |  |  | -  MV=99% | - MV=100% | - MV=100% |
| Pramipexole (salt) |  |  |  | - MV=90% | - MV=89% | - MV=89% |  |  |  | - MV=89% | - MV=89% | - MV=89% |
| Rasagilin |  |  |  | - MV=88% | - MV=75% | - MV=75% |  |  |  | - MV=87% | - MV=75% | - MV=75% |
| Ropinirol |  |  |  | - MV=96% | - MV=93% | - MV=93% |  |  |  | - MV=96% | - MV=93% | - MV=93% |
| Rotigotin |  |  |  | - MV=98% | - MV=96% | - MV=96% |  |  |  | - MV=98% | - MV=96% | - MV=96% |
| Safinamid |  |  |  | - MV=100% | - MV=100% | - MV=100% |  |  |  | - MV=100% | - MV=100% | - MV=100% |
| Selegilinoral |  |  |  | - MV=99 | - MV=96% | - MV=96% |  |  |  | - MV=99% | - MV=96% | - MV=96% |
| Tolcapon |  |  |  | - MV=100% | - MV=100% | - MV=100% |  |  |  | - MV=100% | - MV=100% | - MV=100% |
| **Imaging Features** | | | | | | | | | | | | |
| Brain total |  | + |  |  | + |  |  | + |  |  | + |  |
| Cerebrum right hemisphere |  | + |  |  | + |  |  | + |  |  | + |  |
| Cerebrum left hemisphere |  | - r>0.80 |  |  | - r>0.80 |  |  | - r>0.80 |  |  | - r>0.80 |  |
| T1 hypointensities |  | + |  |  | + |  |  | + |  |  | + |  |
| TIV |  | + |  |  | + |  |  | - r>0.80 |  |  | + |  |
| Midbrain |  | - r>0.80 |  |  | - r>0.80 |  |  | + |  |  | - r>0.80 |  |
| Midbrain-to-Pons-Ratio |  | + |  |  | + |  |  | + |  |  | - r>0.80 |  |
| Ponss |  | + |  |  | - r>0.80 |  |  | + |  |  | + |  |
| SCPP |  | + |  |  | + |  |  | + |  |  | - r>0.80 |  |
| Cerebellum total |  | - r>0.80 |  |  | - r>0.80 |  |  | + |  |  | - r>0.80 |  |
| Fourth ventricle |  | + |  |  | + |  |  | + |  |  | + |  |
| Cerebellum left hemisphere |  | - r>0.80 |  |  | - r>0.80 |  |  | - r>0.80 |  |  | - r>0.80 |  |
| Cerebellum right hemisphere |  | - r>0.80 |  |  | - r>0.80 |  |  | - r>0.80 |  |  | - r>0.80 |  |
| Frontal lobe left |  | - r>0.80 |  |  | - r>0.80 |  |  | + |  |  | - r>0.80 |  |
| Frontal lobe right |  | - r>0.80 |  |  | - r>0.80 |  |  | - r>0.80 |  |  | - r>0.80 |  |
| Gyrus cinguli left |  | - r>0.80 |  |  | + |  |  | + |  |  | - r>0.80 |  |
| Gyrus cinguli right |  | + |  |  | + |  |  | + |  |  | - r>0.80 |  |
| Insula left |  | - r>0.80 |  |  | - r>0.80 |  |  | + |  |  | - r>0.80 |  |
| Insula right |  | - r>0.80 |  |  | - r>0.80 |  |  | - r>0.80 |  |  | - r>0.80 |  |
| Occipital lobe left |  | - r>0.80 |  |  | - r>0.80 |  |  | + |  |  | - r>0.80 |  |
| Occipital lobe right |  | - r>0.80 |  |  | - r>0.80 |  |  | - r>0.80 |  |  | - r>0.80 |  |
| Parietal lobe left |  | - r>0.80 |  |  | - r>0.80 |  |  | - r>0.80 |  |  | - r>0.80 |  |
| Parietal lobe right |  | - r>0.80 |  |  | - r>0.80 |  |  | - r>0.80 |  |  | - r>0.80 |  |
| Temporal lobe left |  | - r>0.80 |  |  | - r>0.80 |  |  | - r>0.80 |  |  | - r>0.80 |  |
| Temporal lobe right |  | - r>0.80 |  |  | - r>0.80 |  |  | + |  |  | - r>0.80 |  |
| Hippocampus left |  | + |  |  | + |  |  | + |  |  | + |  |
| Hippocampus right |  | - r>0.80 |  |  | - r>0.80 |  |  | - r>0.80 |  |  | - r>0.80 |  |
| Lateral ventricle left |  | + |  |  | + |  |  | + |  |  | + |  |
| Lateral ventricle right |  | - r>0.80 |  |  | - r>0.80 |  |  | - r>0.80 |  |  | - r>0.80 |  |
| Supratentorial ventricle |  | - r>0.80 |  |  | - r>0.80 |  |  | - r>0.80 |  |  | - r>0.80 |  |
| Third ventricle |  | - r>0.80 |  |  | + |  |  | + |  |  | - r>0.80 |  |
| Total GM |  | - r>0.80 |  |  | - r>0.80 |  |  | - r>0.80 |  |  | - r>0.80 |  |
| Total GM cortical |  | - r>0.80 |  |  | - r>0.80 |  |  | - r>0.80 |  |  | - r>0.80 |  |
| Total WM |  | - r>0.80 |  |  | - r>0.80 |  |  | - r>0.80 |  |  | + |  |
| **Laboratory values** | | | | | | | | | | | | |
| Amyloid beta 1-42 (Liquor) |  |  |  | - MV=48% | + | + |  |  |  | - MV=51% | + | + |
| Erythrozyten (Liquor) |  |  |  | - MV=99% | - MV=100% | - MV=100% |  |  |  | - MV=99% | - MV=100% | - MV=100% |
| Glucose (Liquor) |  |  |  | - MV=51% | + | + |  |  |  | - MV=53% | + | + |
| IgG (Serum) |  |  |  | - MV=85% | - MV=93% | - MV=93% |  |  |  | - MV=86% | - MV=93% | - MV=93% |
| Leukocytes (Liquor) |  |  |  | - MV=99% | - MV=100% | - MV=100% |  |  |  | - MV=100% | - MV=100% | - MV=100% |
| NFL (Liquor) |  |  |  | - MV=74% | + | + |  |  |  | - MV=76% | + | + |
| Protein (Liquor) |  |  |  | - MV=47% | + | + |  |  |  | - MV=50% | + | + |
| p-Tau (Liquor) |  |  |  | - MV=41% | + | + |  |  |  | - MV=44% | + | + |
| **Single Nucleotide Polymorphisms (SNPs)** | | | | | | | | | | | | |
| SNP: APOE |  |  |  | - MV=49% | - MV=91% | - MV=91% |  |  |  | - MV=47% | - MV=91% | - MV=91% |
| SNP: p. E365K_rs2230288 |  |  |  | - MV=67% | - MV=100% | - MV=100% |  |  |  | - MV=65% | - MV=100% | - MV=100% |
| SNP: p.L444P_rs35095275_rs421016 |  |  |  | - MV=69% | - MV=100% | - MV=100% |  |  |  | - MV=67% | - MV=100% | - MV=100% |
| SNP: p.T336S |  |  |  | + | - MV=78% | + |  |  |  | + | - MV=78% | - MV=78% |
| SNP: p.T408M_rs75548401 |  |  |  | - r>0.80 | - MV=80% | - r>0.80 |  |  |  | - r>0.80 | - MV=80% | - MV=80% |
| SNP: rs34637584 |  |  |  | + | - MV=75% | + |  |  |  | + | - MV=75% | - MV=75 |
| SNP: rs35870237_LRRK2_p.I2020T |  |  |  | + | - MV=76% | + |  |  |  | + | - MV=76% | - MV=76% |
| SNP: rs76763715 |  |  |  | - r>0.80 | - MV=74% | - r>0.80 |  |  |  | - r>0.80 | - MV=74% | - MV=74% |

**Supplementary Table 1**. **Overview of features** considered for analysis (+) or excluded due various reasons (missing value, MV, above the defined threshold, correlation r>0.80 with other variables in the model, excluded by investigators decided for subitems of assessments of the minimal data set); only features potentially included into the analysis are reported for each data set, empty fields indicate features not considered for analysis.

| **Method** | **Mean Absolute Error** | **R-squared Score** | **Mean Squared Logarithmic Error** | **Explained Variance Score** |
| --- | --- | --- | --- | --- |
| KNN | 3.064 | 0.564 | 0.14 | 0.577 |
| Iterative Imputation | 2.484 | 0.669 | 0.104 | 0.677 |
| MissForest | 2.835 | 0.699 | 0.127 | 0.707 |
| MICE | 2.484 | 0.633 | 0.108 | 0.646 |

**Supplementary Table 2. Comparison of imputation methods.** Performance metrics for MICE, MissForest, and KNN imputation are shown, along with results from multiple imputation iterations using MICE. Multiple imputation slightly improved outcomes by averaging across repeated imputations.

| **Model** | **Endpoint** | **Featureset** | **Visits** | **Balanced accuracy** | **ROC-AUC** | **F1** | **Precision** | **Sensitivity** | **Specificity** | **Youden Index** |
| --- | --- | --- | --- | --- | --- | --- | --- | --- | --- | --- |
| Logistic Regression | MCI | Minimal | M0 | 0.62 | 0.7 (± 0.04) | 0.49 | 0.58 | 0.42 | 0.81 | 0.23 |
| Random Forest | MCI | Minimal | M0 | 0.62 | 0.66 (± 0.04) | 0.5 | 0.57 | 0.44 | 0.81 | 0.25 |
| EBM | MCI | Minimal | M0 | 0.62 | 0.66 (± 0.04) | 0.54 | 0.49 | 0.6 | 0.64 | 0.24 |
| MLP | MCI | Minimal | M0 | 0.52 | 0.62 (± 0.04) | 0.17 | 0.53 | 0.1 | 0.95 | 0.05 |
| XGBoost | MCI | Minimal | M0 | 0.63 | 0.68 (± 0.04) | 0.52 | 0.56 | 0.49 | 0.77 | 0.26 |
| Logistic Regression | MCI | Extended | M0 | 0.58 | 0.63 (± 0.05) | 0.42 | 0.44 | 0.41 | 0.74 | 0.15 |
| Random Forest | MCI | Extended | M0 | 0.62 | 0.62 (± 0.05) | 0.49 | 0.48 | 0.5 | 0.73 | 0.23 |
| EBM | MCI | Extended | M0 | 0.58 | 0.6 (± 0.05) | 0.46 | 0.42 | 0.5 | 0.67 | 0.17 |
| MLP | MCI | Extended | M0 | 0.56 | 0.6 (± 0.05) | 0.43 | 0.39 | 0.48 | 0.63 | 0.11 |
| XGBoost | MCI | Extended | M0 | 0.6 | 0.63 (± 0.05) | 0.47 | 0.46 | 0.48 | 0.72 | 0.2 |
| Logistic Regression | MCI | Minimal | M0, M12 | 0.65 | 0.72 (± 0.04) | 0.5 | 0.73 | 0.38 | 0.92 | 0.3 |
| Random Forest | MCI | Minimal | M0, M12 | 0.65 | 0.72 (± 0.04) | 0.52 | 0.7 | 0.41 | 0.9 | 0.31 |
| EBM | MCI | Minimal | M0, M12 | 0.61 | 0.69 (± 0.04) | 0.55 | 0.48 | 0.65 | 0.58 | 0.23 |
| MLP | MCI | Minimal | M0, M12 | 0.67 | 0.73 (± 0.04) | 0.58 | 0.61 | 0.55 | 0.79 | 0.34 |
| XGBoost | MCI | Minimal | M0, M12 | 0.66 | 0.7 (± 0.04) | 0.54 | 0.63 | 0.47 | 0.84 | 0.31 |
| Logistic Regression | MCI | Extended | M0, M12 | 0.6 | 0.63 (± 0.04) | 0.41 | 0.54 | 0.33 | 0.87 | 0.2 |
| Random Forest | MCI | Extended | M0, M12 | 0.63 | 0.68 (± 0.04) | 0.46 | 0.59 | 0.38 | 0.88 | 0.26 |
| EBM | MCI | Extended | M0, M12 | 0.58 | 0.65 (± 0.04) | 0.44 | 0.43 | 0.45 | 0.71 | 0.16 |
| MLP | MCI | Extended | M0, M12 | 0.57 | 0.62 (± 0.05) | 0.4 | 0.43 | 0.38 | 0.76 | 0.14 |
| XGBoost | MCI | Extended | M0, M12 | 0.59 | 0.67 (± 0.05) | 0.42 | 0.48 | 0.38 | 0.8 | 0.18 |
| Logistic Regression | MCI | Minimal | M0, M12, M24 | 0.63 | 0.76 (± 0.03) | 0.48 | 0.65 | 0.39 | 0.87 | 0.26 |
| Random Forest | MCI | Minimal | M0, M12, M24 | 0.69 | 0.77 (± 0.03) | 0.6 | 0.64 | 0.56 | 0.81 | 0.37 |
| EBM | MCI | Minimal | M0, M12, M24 | 0.69 | 0.73 (± 0.04) | 0.63 | 0.56 | 0.72 | 0.66 | 0.38 |
| MLP | MCI | Minimal | M0, M12, M24 | 0.63 | 0.71 (± 0.04) | 0.52 | 0.55 | 0.49 | 0.76 | 0.25 |
| XGBoost | MCI | Minimal | M0, M12, M24 | 0.67 | 0.75 (± 0.04) | 0.56 | 0.66 | 0.49 | 0.85 | 0.34 |
| Logistic Regression | MCI | Extended | M0, M12, M24 | 0.59 | 0.69 (± 0.04) | 0.42 | 0.5 | 0.36 | 0.82 | 0.18 |
| Random Forest | MCI | Extended | M0, M12, M24 | 0.66 | 0.71 (± 0.04) | 0.52 | 0.62 | 0.45 | 0.87 | 0.32 |
| EBM | MCI | Extended | M0, M12, M24 | 0.64 | 0.72 (± 0.04) | 0.5 | 0.55 | 0.47 | 0.82 | 0.29 |
| MLP | MCI | Extended | M0, M12, M24 | 0.59 | 0.62 (± 0.04) | 0.43 | 0.49 | 0.38 | 0.81 | 0.19 |
| XGBoost | MCI | Extended | M0, M12, M24 | 0.66 | 0.73 (± 0.04) | 0.51 | 0.67 | 0.41 | 0.9 | 0.31 |
| Logistic Regression | MCI | Minimal | M0, M12, M24, M36 | 0.62 | 0.72 (± 0.04) | 0.47 | 0.61 | 0.39 | 0.85 | 0.24 |
| Random Forest | MCI | Minimal | M0, M12, M24, M36 | 0.68 | 0.77 (± 0.03) | 0.58 | 0.66 | 0.53 | 0.84 | 0.37 |
| EBM | MCI | Minimal | M0, M12, M24, M36 | 0.68 | 0.73 (± 0.04) | 0.61 | 0.56 | 0.68 | 0.69 | 0.37 |
| MLP | MCI | Minimal | M0, M12, M24, M36 | 0.65 | 0.74 (± 0.03) | 0.55 | 0.58 | 0.53 | 0.77 | 0.3 |
| Logistic Regression | MCI | Extended | M0, M12, M24, M36 | 0.56 | 0.66 (± 0.04) | 0.3 | 0.55 | 0.21 | 0.92 | 0.13 |
| Random Forest | MCI | Extended | M0, M12, M24, M36 | 0.62 | 0.73 (± 0.04) | 0.46 | 0.58 | 0.38 | 0.87 | 0.25 |
| EBM | MCI | Extended | M0, M12, M24, M36 | 0.66 | 0.69 (± 0.04) | 0.54 | 0.55 | 0.53 | 0.79 | 0.32 |
| MLP | MCI | Extended | M0, M12, M24, M36 | 0.55 | 0.62 (± 0.04) | 0.34 | 0.44 | 0.28 | 0.83 | 0.11 |

**Supplementary Table 3a: Model performances of the clinicodemographic minimal and multimodal extended datasets for the prediction of the clinical endpoint MCI using data from different visits.** ROC-AUC scores have been calculated using bootstrapping with 1000 samples. Youden Index is defined as sensitivity + specificity – 1.

| **Model** | **Endpoint** | **Featureset** | **Visits** | **Balanced  accuracy** | **ROC-AUC** | **F1** | **Precision** | **Sensitivity** | **Specificity** | **Youden Index** |
| --- | --- | --- | --- | --- | --- | --- | --- | --- | --- | --- |
| Logistic Regression | PI | Minimal | M0 | 0.59 | 0.63 (± 0.05) | 0.34 | 0.24 | 0.62 | 0.56 | 0.18 |
| Random Forest | PI | Minimal | M0 | 0.6 | 0.64 (± 0.05) | 0.35 | 0.25 | 0.58 | 0.63 | 0.21 |
| EBM | PI | Minimal | M0 | 0.61 | 0.63 (± 0.05) | 0.36 | 0.25 | 0.67 | 0.55 | 0.22 |
| MLP | PI | Minimal | M0 | 0.5 | 0.54 (± 0.04) | 0 | NaN | 0 | 1 | 0 |
| XGBoost | PI | Minimal | M0 | 0.57 | 0.61 (± 0.05) | 0.31 | 0.24 | 0.44 | 0.69 | 0.13 |
| Logistic Regression | PI | Extended | M0 | 0.63 | 0.65 (± 0.05) | 0.35 | 0.23 | 0.69 | 0.56 | 0.25 |
| Random Forest | PI | Extended | M0 | 0.6 | 0.65 (± 0.06) | 0.32 | 0.38 | 0.28 | 0.91 | 0.19 |
| EBM | PI | Extended | M0 | 0.61 | 0.65 (± 0.05) | 0.35 | 0.29 | 0.44 | 0.79 | 0.23 |
| MLP | PI | Extended | M0 | 0.58 | 0.59 (± 0.06) | 0.31 | 0.23 | 0.47 | 0.69 | 0.16 |
| XGBoost | PI | Extended | M0 | 0.57 | 0.64 (± 0.06) | 0.29 | 0.25 | 0.34 | 0.8 | 0.14 |
| Logistic Regression | PI | Minimal | M0, M12 | 0.55 | 0.61 (± 0.05) | 0.32 | 0.21 | 0.64 | 0.46 | 0.1 |
| Random Forest | PI | Minimal | M0, M12 | 0.6 | 0.64 (± 0.05) | 0.35 | 0.26 | 0.53 | 0.66 | 0.19 |
| EBM | PI | Minimal | M0, M12 | 0.59 | 0.65 (± 0.05) | 0.34 | 0.3 | 0.4 | 0.79 | 0.19 |
| MLP | PI | Minimal | M0, M12 | 0.52 | 0.59 (± 0.05) | 0.11 | 0.33 | 0.07 | 0.97 | 0.04 |
| XGBoost | PI | Minimal | M0, M12 | 0.57 | 0.61 (± 0.05) | 0.32 | 0.26 | 0.4 | 0.75 | 0.15 |
| Logistic Regression | PI | Extended | M0, M12 | 0.56 | 0.65 (± 0.05) | 0.27 | 0.24 | 0.31 | 0.81 | 0.12 |
| Random Forest | PI | Extended | M0, M12 | 0.55 | 0.68 (± 0.05) | 0.24 | 0.32 | 0.19 | 0.92 | 0.11 |
| EBM | PI | Extended | M0, M12 | 0.54 | 0.65 (± 0.05) | 0.2 | 0.29 | 0.16 | 0.93 | 0.09 |
| MLP | PI | Extended | M0, M12 | 0.57 | 0.64 (± 0.06) | 0.27 | 0.29 | 0.25 | 0.88 | 0.13 |
| XGBoost | PI | Extended | M0, M12 | 0.59 | 0.71 (± 0.05) | 0.31 | 0.35 | 0.28 | 0.9 | 0.18 |
| Logistic Regression | PI | Minimal | M0, M12, M24 | 0.5 | 0.58 (± 0.04) | 0.04 | 0.25 | 0.02 | 0.99 | 0.01 |
| Random Forest | PI | Minimal | M0, M12, M24 | 0.52 | 0.56 (± 0.05) | 0.11 | 0.33 | 0.07 | 0.97 | 0.04 |
| EBM | PI | Minimal | M0, M12, M24 | 0.52 | 0.63 (± 0.05) | 0.08 | 0.5 | 0.04 | 0.99 | 0.03 |
| MLP | PI | Minimal | M0, M12, M24 | 0.51 | 0.46 (± 0.05) | 0.16 | 0.19 | 0.13 | 0.88 | 0.01 |
| XGBoost | PI | Minimal | M0, M12, M24 | 0.53 | 0.61 (± 0.05) | 0.12 | 0.6 | 0.07 | 0.99 | 0.06 |
| Logistic Regression | PI | Extended | M0, M12, M24 | 0.52 | 0.72 (± 0.05) | 0.11 | 0.33 | 0.06 | 0.98 | 0.04 |
| Random Forest | PI | Extended | M0, M12, M24 | 0.55 | 0.7 (± 0.05) | 0.2 | 0.44 | 0.12 | 0.97 | 0.09 |
| EBM | PI | Extended | M0, M12, M24 | 0.56 | 0.74 (± 0.05) | 0.22 | 1 | 0.12 | 1 | 0.12 |
| MLP | PI | Extended | M0, M12, M24 | 0.55 | 0.73 (± 0.04) | 0.21 | 0.31 | 0.16 | 0.93 | 0.09 |
| XGBoost | PI | Extended | M0, M12, M24 | 0.59 | 0.71 (± 0.05) | 0.31 | 0.35 | 0.28 | 0.9 | 0.18 |
| Logistic Regression | PI | Minimal | M0, M12, M24, M36 | 0.64 | 0.76 (± 0.03) | 0.51 | 0.66 | 0.41 | 0.87 | 0.28 |
| Random Forest | PI | Minimal | M0, M12, M24, M36 | 0.69 | 0.79 (± 0.03) | 0.61 | 0.65 | 0.57 | 0.81 | 0.38 |
| EBM | PI | Minimal | M0, M12, M24, M36 | 0.69 | 0.73 (± 0.04) | 0.63 | 0.56 | 0.72 | 0.66 | 0.38 |
| MLP | PI | Minimal | M0, M12, M24, M36 | 0.61 | 0.76 (± 0.03) | 0.44 | 0.67 | 0.33 | 0.9 | 0.23 |
| XGBoost | PI | Minimal | M0, M12, M24, M36 | 0.63 | 0.74 (± 0.03) | 0.51 | 0.59 | 0.45 | 0.81 | 0.26 |
| Logistic Regression | PI | Extended | M0, M12, M24, M36 | 0.55 | 0.66 (± 0.04) | 0.28 | 0.52 | 0.19 | 0.92 | 0.11 |
| Random Forest | PI | Extended | M0, M12, M24, M36 | 0.61 | 0.69 (± 0.04) | 0.43 | 0.57 | 0.34 | 0.88 | 0.22 |
| EBM | PI | Extended | M0, M12, M24, M36 | 0.65 | 0.71 (± 0.04) | 0.52 | 0.56 | 0.48 | 0.82 | 0.3 |
| MLP | PI | Extended | M0, M12, M24, M36 | 0.54 | 0.63 (± 0.05) | 0.33 | 0.41 | 0.28 | 0.81 | 0.09 |
| XGBoost | PI | Extended | M0, M12, M24, M36 | 0.6 | 0.68 (± 0.04) | 0.42 | 0.51 | 0.36 | 0.83 | 0.19 |

**Supplementary Table 3b Model performances of the clinicodemographic minimal and multimodal extended datasets for the prediction of the clinical endpoint PI using data from different visits.** ROC-AUC scores have been calculated using bootstrapping with 1000 samples. Youden Index is defined as sensitivity + specificity – 1.

| **Model** | **Endpoint** | **Featureset** | **Hyperparameter Optimization** | **Feature engineering** | **Sample Weight (P/N)** | **Balanced accuracy** | **ROC AUC** | **F1** | **Precision** | **Sensitivity** | **Specificity** | **Youden Index** |
| --- | --- | --- | --- | --- | --- | --- | --- | --- | --- | --- | --- | --- |
| EBM | MCI | Minimal | False | True | 1:1 | 0.62 | 0.67 (± 0.04) | 0.55 | 0.5 | 0.61 | 0.64 | 0.25 |
| EBM | MCI | Extended | False | False | 1:1 | 0.63 | 0.67 (± 0.04) | 0.55 | 0.51 | 0.6 | 0.67 | 0.27 |

**Supplementary Table 4a EBM model performances of the clinicodemographic minimal and multimodal extended datasets for the prediction of the clinical endpoint MCI.** ROC-AUC scores have been calculated using bootstrapping with 1000 samples. Youden Index is defined as sensitivity + specificity – 1.

| **Model** | **Endpoint** | **Featureset** | **Hyperparameter Optimization** | **Feature engineering** | **Sample Weight (P/N)** | **Balanced accuracy** | **ROC AUC** | **F1** | **Precision** | **Sensitivity** | **Specificity** | **Youden Index** |
| --- | --- | --- | --- | --- | --- | --- | --- | --- | --- | --- | --- | --- |
| EBM | PI | Minimal | False | True | 10:1 | 0.61 | 0.62 (± 0.05) | 0.36 | 0.24 | 0.73 | 0.48 | 0.21 |
| EBM | PI | Minimal | False | True | 5:1 | 0.56 | 0.62 (± 0.05) | 0.29 | 0.28 | 0.29 | 0.84 | 0.13 |
| EBM | PI | Minimal | False | True | 8:1 | 0.6 | 0.62 (± 0.05) | 0.35 | 0.24 | 0.64 | 0.55 | 0.19 |
| EBM | PI | Minimal | False | True | 6.6:1 | 0.61 | 0.62 (± 0.05) | 0.36 | 0.28 | 0.53 | 0.69 | 0.22 |
| EBM | PI | Minimal | False | True | 1:1 | 0.5 | 0.61 (± 0.05) | 0 | NaN | 0 | 1 | 0 |
| EBM | PI | Minimal | False | False | 10:1 | 0.61 | 0.62 (± 0.05) | 0.36 | 0.24 | 0.73 | 0.48 | 0.21 |
| EBM | PI | Minimal | False | False | 5:1 | 0.56 | 0.62 (± 0.05) | 0.29 | 0.28 | 0.29 | 0.84 | 0.13 |
| EBM | PI | Minimal | False | False | 8:1 | 0.6 | 0.62 (± 0.05) | 0.35 | 0.24 | 0.64 | 0.55 | 0.19 |
| EBM | PI | Minimal | False | False | 6.6:1 | 0.6 | 0.62 (± 0.05) | 0.35 | 0.27 | 0.51 | 0.69 | 0.2 |
| EBM | PI | Minimal | False | False | 1:1 | 0.5 | 0.59 (± 0.05) | 0 | NaN | 0 | 1 | 0 |
| EBM | PI | Minimal | True | True | 10:1 | 0.61 | 0.62 (± 0.05) | 0.36 | 0.24 | 0.73 | 0.48 | 0.21 |
| EBM | PI | Minimal | True | True | 5:1 | 0.57 | 0.65 (± 0.05) | 0.29 | 0.32 | 0.27 | 0.87 | 0.14 |
| EBM | PI | Minimal | True | True | 8:1 | 0.6 | 0.62 (± 0.05) | 0.36 | 0.26 | 0.56 | 0.65 | 0.21 |
| EBM | PI | Minimal | True | True | 6.6:1 | 0.6 | 0.62 (± 0.05) | 0.35 | 0.28 | 0.47 | 0.74 | 0.21 |
| EBM | PI | Minimal | True | True | 1:1 | 0.5 | 0.63 (± 0.05) | 0 | NaN | 0 | 1 | 0 |
| EBM | PI | Minimal | True | False | 10:1 | 0.61 | 0.63 (± 0.05) | 0.36 | 0.24 | 0.73 | 0.48 | 0.21 |
| EBM | PI | Minimal | True | False | 5:1 | 0.6 | 0.65 (± 0.05) | 0.34 | 0.33 | 0.36 | 0.84 | 0.2 |
| EBM | PI | Minimal | True | False | 8:1 | 0.58 | 0.63 (± 0.05) | 0.34 | 0.24 | 0.56 | 0.61 | 0.17 |
| EBM | PI | Minimal | True | False | 6.6:1 | 0.6 | 0.64 (± 0.05) | 0.35 | 0.27 | 0.49 | 0.71 | 0.2 |
| EBM | PI | Minimal | True | False | 1:1 | 0.5 | 0.65 (± 0.04) | 0 | NaN | 0 | 1 | 0 |
| EBM | PI | Extended | False | True | 10:1 | 0.58 | 0.63 (± 0.06) | 0.31 | 0.2 | 0.72 | 0.44 | 0.16 |
| EBM | PI | Extended | False | True | 5:1 | 0.52 | 0.65 (± 0.05) | 0.18 | 0.21 | 0.16 | 0.89 | 0.05 |
| EBM | PI | Extended | False | True | 8:1 | 0.6 | 0.64 (± 0.05) | 0.33 | 0.23 | 0.59 | 0.61 | 0.2 |
| EBM | PI | Extended | False | True | 6.6:1 | 0.57 | 0.65 (± 0.05) | 0.29 | 0.22 | 0.44 | 0.7 | 0.14 |
| EBM | PI | Extended | False | True | 1:1 | 0.5 | 0.65 (± 0.05) | 0 | NaN | 0 | 1 | 0 |
| EBM | PI | Extended | False | False | 10:1 | 0.62 | 0.64 (± 0.05) | 0.34 | 0.21 | 0.81 | 0.42 | 0.23 |
| EBM | PI | Extended | False | False | 5:1 | 0.57 | 0.65 (± 0.05) | 0.27 | 0.35 | 0.22 | 0.92 | 0.14 |
| EBM | PI | Extended | False | False | 8:1 | 0.6 | 0.65 (± 0.06) | 0.33 | 0.22 | 0.62 | 0.58 | 0.2 |
| EBM | PI | Extended | False | False | 6.6:1 | 0.57 | 0.65 (± 0.05) | 0.3 | 0.23 | 0.41 | 0.74 | 0.15 |
| EBM | PI | Extended | False | False | 1:1 | 0.57 | 0.65 (± 0.05) | 0.3 | 0.23 | 0.41 | 0.74 | 0.15 |
| EBM | PI | Extended | True | True | 10:1 | 0.64 | 0.63 (± 0.05) | 0.36 | 0.25 | 0.69 | 0.6 | 0.29 |
| EBM | PI | Extended | True | True | 5:1 | 0.54 | 0.66 (± 0.06) | 0.21 | 0.31 | 0.16 | 0.93 | 0.09 |
| EBM | PI | Extended | True | True | 8:1 | 0.64 | 0.64 (± 0.06) | 0.37 | 0.26 | 0.59 | 0.68 | 0.27 |
| EBM | PI | Extended | True | True | 6.6:1 | 0.58 | 0.64 (± 0.06) | 0.31 | 0.28 | 0.34 | 0.83 | 0.17 |
| EBM | PI | Extended | True | True | 1:1 | 0.5 | 0.66 (± 0.05) | 0 | NaN | 0 | 1 | 0 |
| EBM | PI | Extended | True | False | 10:1 | 0.49 | 0.58 (± 0.06) | 0.27 | 0.16 | 0.97 | 0.02 | -0.01 |
| EBM | PI | Extended | True | False | 5:1 | 0.49 | 0.58 (± 0.06) | 0.27 | 0.16 | 0.97 | 0.02 | -0.01 |
| EBM | PI | Extended | True | False | 8:1 | 0.65 | 0.63 (± 0.06) | 0.37 | 0.26 | 0.66 | 0.64 | 0.3 |
| EBM | PI | Extended | True | False | 6.6:1 | 0.6 | 0.64 (± 0.06) | 0.33 | 0.28 | 0.41 | 0.8 | 0.21 |
| EBM | PI | Extended | True | False | 1:1 | 0.5 | 0.66 (± 0.05) | 0 | NaN | 0 | 1 | 0 |

**Supplementary Table 4b EBM model performances of the clinicodemographic minimal and multimodal extended datasets for the prediction of the clinical endpoint PI.** ROC-AUC scores have been calculated using bootstrapping with 1000 samples. Youden Index is defined as sensitivity + specificity – 1.

| **Model** | **Endpoint** | **Feature set** | **Imaging** | **Split size** | **Sample Weight (P/N)** | **Balanced accuracy** | **ROC AUC** | **F1** | **Precision** | **Sensitivity** | **Specificity** | **Youden Index** |
| --- | --- | --- | --- | --- | --- | --- | --- | --- | --- | --- | --- | --- |
| EBM | MCI | Minimal | No | 0.2 | 10:1 | 0.6 | 0.72 (± 0.13) | 0.5 | 0.37 | 0.81 | 0.4 | 0.21 |
| EBM | MCI | Minimal | No | 0.2 | 5:1 | 0.62 | 0.73 (± 0.08) | 0.51 | 0.39 | 0.75 | 0.49 | 0.24 |
| EBM | MCI | Minimal | No | 0.2 | 8:1 | 0.6 | 0.72 (± 0.13) | 0.5 | 0.37 | 0.81 | 0.4 | 0.21 |
| EBM | MCI | Minimal | No | 0.2 | 6.6:1 | 0.6 | 0.72 (± 0.13) | 0.5 | 0.37 | 0.81 | 0.4 | 0.21 |
| EBM | MCI | Minimal | No | 0.2 | 1:1 | 0.6 | 0.72 (± 0.13) | 0.5 | 0.37 | 0.81 | 0.4 | 0.21 |
| EBM | MCI | Extended | No | 0.2 | 10:1 | 0.62 | 0.69 (± 0.13) | 0.52 | 0.4 | 0.78 | 0.45 | 0.23 |
| EBM | MCI | Extended | No | 0.2 | 5:1 | 0.63 | 0.7 (± 0.12) | 0.51 | 0.46 | 0.61 | 0.66 | 0.27 |
| EBM | MCI | Extended | No | 0.2 | 8:1 | 0.6 | 0.69 (± 0.13) | 0.48 | 0.39 | 0.69 | 0.51 | 0.2 |
| EBM | MCI | Extended | No | 0.2 | 6.6:1 | 0.62 | 0.7 (± 0.12) | 0.5 | 0.42 | 0.66 | 0.57 | 0.23 |
| EBM | MCI | Extended | No | 0.2 | 1:1 | 0.61 | 0.73 (± 0.11) | 0.4 | 0.59 | 0.34 | 0.89 | 0.23 |
| EBM | MCI | Minimal | No | 0.4 | 10:1 | 0.62 | 0.73 (± 0.08) | 0.51 | 0.39 | 0.75 | 0.49 | 0.24 |
| EBM | MCI | Minimal | No | 0.4 | 5:1 | 0.65 | 0.73 (± 0.08) | 0.53 | 0.54 | 0.56 | 0.75 | 0.31 |
| EBM | MCI | Minimal | No | 0.4 | 8:1 | 0.64 | 0.74 (± 0.08) | 0.52 | 0.48 | 0.62 | 0.66 | 0.28 |
| EBM | MCI | Minimal | No | 0.4 | 6.6:1 | 0.65 | 0.73 (± 0.08) | 0.53 | 0.54 | 0.56 | 0.75 | 0.31 |
| EBM | MCI | Minimal | No | 0.4 | 1:1 | 0.64 | 0.74 (± 0.09) | 0.52 | 0.53 | 0.59 | 0.7 | 0.29 |
| EBM | MCI | Extended | No | 0.4 | 10:1 | 0.63 | 0.69 (± 0.09) | 0.52 | 0.42 | 0.73 | 0.53 | 0.26 |
| EBM | MCI | Extended | No | 0.4 | 5:1 | 0.62 | 0.69 (± 0.09) | 0.48 | 0.43 | 0.57 | 0.66 | 0.23 |
| EBM | MCI | Extended | No | 0.4 | 8:1 | 0.63 | 0.69 (± 0.09) | 0.51 | 0.43 | 0.71 | 0.56 | 0.27 |
| EBM | MCI | Extended | No | 0.4 | 6.6:1 | 0.63 | 0.69 (± 0.08) | 0.51 | 0.43 | 0.68 | 0.59 | 0.27 |
| EBM | MCI | Extended | No | 0.4 | 1:1 | 0.61 | 0.72 (± 0.08) | 0.42 | 0.61 | 0.34 | 0.88 | 0.22 |
| EBM | MCI | Minimal | Yes | 0.2 | 10:1 | 0.64 | 0.72 (± 0.12) | 0.52 | 0.41 | 0.74 | 0.53 | 0.27 |
| EBM | MCI | Minimal | Yes | 0.2 | 5:1 | 0.66 | 0.74 (± 0.12) | 0.55 | 0.46 | 0.7 | 0.63 | 0.33 |
| EBM | MCI | Minimal | Yes | 0.2 | 8:1 | 0.65 | 0.73 (± 0.12) | 0.53 | 0.43 | 0.71 | 0.59 | 0.3 |
| EBM | MCI | Minimal | Yes | 0.2 | 6.6:1 | 0.66 | 0.73 (± 0.12) | 0.54 | 0.45 | 0.71 | 0.6 | 0.31 |
| EBM | MCI | Minimal | Yes | 0.2 | 1:1 | 0.65 | 0.74 (± 0.12) | 0.52 | 0.58 | 0.48 | 0.82 | 0.3 |
| EBM | MCI | Extended | Yes | 0.2 | 10:1 | 0.58 | 0.66 (± 0.13) | 0.48 | 0.36 | 0.77 | 0.39 | 0.16 |
| EBM | MCI | Extended | Yes | 0.2 | 5:1 | 0.6 | 0.68 (± 0.13) | 0.46 | 0.4 | 0.57 | 0.62 | 0.19 |
| EBM | MCI | Extended | Yes | 0.2 | 8:1 | 0.61 | 0.67 (± 0.13) | 0.5 | 0.41 | 0.71 | 0.51 | 0.22 |
| EBM | MCI | Extended | Yes | 0.2 | 6.6:1 | 0.62 | 0.67 (± 0.13) | 0.49 | 0.41 | 0.66 | 0.57 | 0.23 |
| EBM | MCI | Extended | Yes | 0.2 | 1:1 | 0.61 | 0.7 (± 0.12) | 0.39 | 0.6 | 0.32 | 0.89 | 0.21 |
| EBM | MCI | Minimal | Yes | 0.4 | 10:1 | 0.67 | 0.74 (± 0.09) | 0.56 | 0.43 | 0.82 | 0.52 | 0.34 |
| EBM | MCI | Minimal | Yes | 0.4 | 5:1 | 0.68 | 0.74 (± 0.09) | 0.56 | 0.46 | 0.73 | 0.62 | 0.35 |
| EBM | MCI | Minimal | Yes | 0.4 | 8:1 | 0.67 | 0.74 (± 0.09) | 0.55 | 0.42 | 0.81 | 0.52 | 0.33 |
| EBM | MCI | Minimal | Yes | 0.4 | 6.6:1 | 0.68 | 0.74 (± 0.09) | 0.57 | 0.45 | 0.79 | 0.58 | 0.37 |
| EBM | MCI | Minimal | Yes | 0.4 | 1:1 | 0.64 | 0.72 (± 0.09) | 0.49 | 0.5 | 0.51 | 0.77 | 0.28 |
| EBM | MCI | Extended | Yes | 0.4 | 10:1 | 0.61 | 0.66 (± 0.09) | 0.49 | 0.4 | 0.69 | 0.53 | 0.22 |
| EBM | MCI | Extended | Yes | 0.4 | 5:1 | 0.6 | 0.68 (± 0.09) | 0.46 | 0.41 | 0.55 | 0.65 | 0.2 |
| EBM | MCI | Extended | Yes | 0.4 | 8:1 | 0.61 | 0.67 (± 0.09) | 0.48 | 0.41 | 0.64 | 0.57 | 0.21 |
| EBM | MCI | Extended | Yes | 0.4 | 6.6:1 | 0.6 | 0.67 (± 0.09) | 0.46 | 0.4 | 0.6 | 0.59 | 0.19 |
| EBM | MCI | Extended | Yes | 0.4 | 1:1 | 0.58 | 0.71 (± 0.08) | 0.34 | 0.54 | 0.26 | 0.89 | 0.15 |

**Supplementary Table 5a EBM model performances of the imaging cohort, assessed with and without imaging data, in predicting the clinical endpoint MCI.** ROC-AUC scores have been calculated using bootstrapping with 1000 samples. Youden Index is defined as sensitivity + specificity – 1.

| **Model** | **Endpoint** | **Feature set** | **Imaging** | **Split size** | **Sample Weight (P/N)** | **Balanced accuracy** | **ROC AUC** | **F1** | **Precision** | **Sensitivity** | **Specificity** | **Youden Index** |
| --- | --- | --- | --- | --- | --- | --- | --- | --- | --- | --- | --- | --- |
| EBM | PI | Minimal | No | 0.2 | 10:1 | 0.58 | 0.67 (± 0.16) | 0.28 | 0.21 | 0.52 | 0.65 | 0.17 |
| EBM | PI | Minimal | No | 0.2 | 5:1 | 0.59 | 0.62 (± 0.16) | 0.32 | 0.4 | 0.36 | 0.82 | 0.18 |
| EBM | PI | Minimal | No | 0.2 | 8:1 | 0.59 | 0.66 (± 0.17) | 0.31 | 0.3 | 0.48 | 0.7 | 0.18 |
| EBM | PI | Minimal | No | 0.2 | 6.6:1 | 0.59 | 0.62 (± 0.17) | 0.31 | 0.4 | 0.32 | 0.85 | 0.17 |
| EBM | PI | Minimal | No | 0.2 | 1:1 | 0.56 | 0.59 (± 0.15) | 0.27 | 0.34 | 0.34 | 0.77 | 0.11 |
| EBM | PI | Extended | No | 0.2 | 10:1 | 0.53 | 0.64 (± 0.16) | 0.13 | 0.19 | 0.15 | 0.91 | 0.06 |
| EBM | PI | Extended | No | 0.2 | 5:1 | 0.51 | 0.63 (± 0.16) | 0.07 | 0.14 | 0.06 | 0.95 | 0.01 |
| EBM | PI | Extended | No | 0.2 | 8:1 | 0.51 | 0.63 (± 0.16) | 0.09 | 0.15 | 0.09 | 0.92 | 0.01 |
| EBM | PI | Extended | No | 0.2 | 6.6:1 | 0.51 | 0.63 (± 0.16) | 0.09 | 0.15 | 0.09 | 0.93 | 0.02 |
| EBM | PI | Extended | No | 0.2 | 1:1 | 0.49 | 0.67 (± 0.16) | 0 | 0 | 0 | 0.98 | -0.02 |
| EBM | PI | Minimal | No | 0.4 | 10:1 | 0.54 | 0.59 (± 0.14) | 0.23 | 0.16 | 0.4 | 0.69 | 0.09 |
| EBM | PI | Minimal | No | 0.4 | 5:1 | 0.55 | 0.64 (± 0.16) | 0.23 | 0.18 | 0.39 | 0.71 | 0.1 |
| EBM | PI | Minimal | No | 0.4 | 8:1 | 0.55 | 0.6 (± 0.13) | 0.21 | 0.15 | 0.36 | 0.74 | 0.1 |
| EBM | PI | Minimal | No | 0.4 | 6.6:1 | 0.59 | 0.64 (± 0.14) | 0.25 | 0.2 | 0.38 | 0.79 | 0.17 |
| EBM | PI | Minimal | No | 0.4 | 1:1 | 0.56 | 0.65 (± 0.16) | 0.24 | 0.2 | 0.41 | 0.7 | 0.11 |
| EBM | PI | Extended | No | 0.4 | 10:1 | 0.52 | 0.67 (± 0.12) | 0.16 | 0.21 | 0.16 | 0.88 | 0.04 |
| EBM | PI | Extended | No | 0.4 | 5:1 | 0.53 | 0.66 (± 0.12) | 0.13 | 0.19 | 0.11 | 0.95 | 0.06 |
| EBM | PI | Extended | No | 0.4 | 8:1 | 0.53 | 0.67 (± 0.12) | 0.15 | 0.18 | 0.15 | 0.91 | 0.06 |
| EBM | PI | Extended | No | 0.4 | 6.6:1 | 0.54 | 0.66 (± 0.12) | 0.16 | 0.21 | 0.15 | 0.93 | 0.08 |
| EBM | PI | Extended | No | 0.4 | 1:1 | 0.51 | 0.69 (± 0.12) | 0.05 | 0.21 | 0.04 | 0.98 | 0.02 |
| EBM | PI | Minimal | Yes | 0.2 | 10:1 | 0.59 | 0.7 (± 0.16) | 0.31 | 0.24 | 0.42 | 0.76 | 0.18 |
| EBM | PI | Minimal | Yes | 0.2 | 5:1 | 0.62 | 0.69 (± 0.17) | 0.35 | 0.34 | 0.42 | 0.82 | 0.24 |
| EBM | PI | Minimal | Yes | 0.2 | 8:1 | 0.6 | 0.71 (± 0.16) | 0.32 | 0.26 | 0.42 | 0.77 | 0.19 |
| EBM | PI | Minimal | Yes | 0.2 | 6.6:1 | 0.61 | 0.7 (± 0.17) | 0.33 | 0.28 | 0.42 | 0.79 | 0.21 |
| EBM | PI | Minimal | Yes | 0.2 | 1:1 | 0.58 | 0.66 (± 0.18) | 0.28 | 0.39 | 0.27 | 0.89 | 0.16 |
| EBM | PI | Extended | Yes | 0.2 | 10:1 | 0.64 | 0.72 (± 0.18) | 0.38 | 0.51 | 0.36 | 0.91 | 0.27 |
| EBM | PI | Extended | Yes | 0.2 | 5:1 | 0.6 | 0.71 (± 0.17) | 0.27 | 0.64 | 0.21 | 0.98 | 0.19 |
| EBM | PI | Extended | Yes | 0.2 | 8:1 | 0.61 | 0.72 (± 0.18) | 0.32 | 0.52 | 0.27 | 0.95 | 0.22 |
| EBM | PI | Extended | Yes | 0.2 | 6.6:1 | 0.61 | 0.71 (± 0.17) | 0.29 | 0.56 | 0.24 | 0.97 | 0.21 |
| EBM | PI | Extended | Yes | 0.2 | 1:1 | 0.5 | 0.75 (± 0.15) | 0 | NaN | 0 | 1 | 0 |
| EBM | PI | Minimal | Yes | 0.4 | 10:1 | 0.63 | 0.68 (± 0.13) | 0.32 | 0.27 | 0.44 | 0.82 | 0.26 |
| EBM | PI | Minimal | Yes | 0.4 | 5:1 | 0.63 | 0.67 (± 0.14) | 0.34 | 0.32 | 0.42 | 0.84 | 0.26 |
| EBM | PI | Minimal | Yes | 0.4 | 8:1 | 0.62 | 0.68 (± 0.13) | 0.31 | 0.26 | 0.42 | 0.81 | 0.23 |
| EBM | PI | Minimal | Yes | 0.4 | 6.6:1 | 0.63 | 0.67 (± 0.14) | 0.34 | 0.29 | 0.44 | 0.82 | 0.26 |
| EBM | PI | Minimal | Yes | 0.4 | 1:1 | 0.57 | 0.65 (± 0.14) | 0.24 | 0.3 | 0.22 | 0.92 | 0.14 |
| EBM | PI | Extended | Yes | 0.4 | 10:1 | 0.58 | 0.74 (± 0.12) | 0.24 | 0.4 | 0.22 | 0.94 | 0.16 |
| EBM | PI | Extended | Yes | 0.4 | 5:1 | 0.58 | 0.74 (± 0.12) | 0.24 | 0.56 | 0.18 | 0.97 | 0.15 |
| EBM | PI | Extended | Yes | 0.4 | 8:1 | 0.56 | 0.74 (± 0.12) | 0.21 | 0.36 | 0.18 | 0.95 | 0.13 |
| EBM | PI | Extended | Yes | 0.4 | 6.6:1 | 0.57 | 0.74 (± 0.12) | 0.23 | 0.5 | 0.18 | 0.96 | 0.14 |
| EBM | PI | Extended | Yes | 0.4 | 1:1 | 0.52 | 0.76 (± 0.11) | 0.08 | 0.5 | 0.05 | 0.99 | 0.04 |

**Supplementary Table 5b EBM model performances of the imaging cohort, assessed with and without imaging data, in predicting the clinical endpoint PI.** ROC-AUC scores have been calculated using bootstrapping with 1000 samples. Youden Index is defined as sensitivity + specificity – 1.
